# Integrative proteogenomic analyses provide novel interpretations of type 1 diabetes risk loci through circulating proteins

**DOI:** 10.1101/2023.12.19.23300201

**Authors:** Tianyuan Lu, Despoina Manousaki, Lei Sun, Andrew D. Paterson

**Affiliations:** Department of Statistical Sciences, Faculty of Arts and Science, University of Toronto, Toronto, ON, Canada; Department of Pediatrics, Faculty of Medicine, Université de Montréal, Montreal, QC, Canada; Department of Biochemistry and Molecular Medicine, Faculty of Medicine, Université de Montréal, Montreal, QC, Canada; Research Center of the Sainte-Justine University Hospital, Université de Montréal, Montreal, QC, Canada; Division of Biostatistics, Dalla Lana School of Public Health, University of Toronto, Toronto, ON, Canada; Division of Epidemiology, Dalla Lana School of Public Health, University of Toronto, Toronto, ON, Canada; Genetics and Genome Biology, The Hospital for Sick Children, Toronto, ON, Canada

## Abstract

Type 1 diabetes (T1D) requires new preventive measures and interventions. Circulating proteins are promising biomarkers and drug targets. Leveraging genome-wide association studies (GWASs) of T1D (18,942 cases and 501,638 controls) and circulating protein abundances (10,708 individuals), the associations between 1,565 circulating proteins and T1D risk were assessed through Mendelian randomization, followed by multiple sensitivity and colocalization analyses, examinations of horizontal pleiotropy, and replications. Genetically increased circulating abundances of CTSH, IL27RA, SIRPG, and PGM1 were associated with an increased risk of T1D, consistently replicated in other cohorts. Bulk tissue and single-cell gene expression profiles revealed strong enrichment of *CTSH, IL27RA*, and SIRPG in immune system-related tissues, and *PGM1* in muscle and liver tissues. Among immune cells, *CTSH* was enriched in B cells and myeloid cells, while *SIRPG* was enriched in T cells and natural killer cells. These proteins warrant exploration as T1D biomarkers or drug targets in relevant tissues.

## Introduction

Type 1 diabetes is an autoimmune disease characterized by the destruction of pancreatic *β* cells ^1-6^, which are responsible for producing insulin. Although traditionally considered a disease of children and adolescents, type 1 diabetes can be diagnosed at any age and can affect a significant proportion of the global population ^3-5^. Despite continuing efforts to develop risk predictors ^7-9^, few effective preventive measures for type 1 diabetes have been implemented in public health practice. Following diagnosis, preserving residual *β* cell function and delaying type 1 diabetes-associated autoimmunity present challenges due to the multifactorial and heterogeneous nature of the disease ^3-5,10^. Consequently, there is an urgent need for new biomarkers and drug targets for type 1 diabetes.

Circulating molecules participate in various biological processes and play essential roles encompassing immune responses, signaling cascades, and regulatory mechanisms ^11-14^. These molecules may be promising biomarkers or drug targets because their abundances are measurable and possibly modifiable. Autoantibodies to insulin, glutamic acid decarboxylase, islet antigen-2, zinc transporter 8, and other circulating proteins have emerged as markers for characterizing type 1 diabetes ^10, 15,16^. Yet, establishing the causal roles of novel proteins is difficult. The feasibility of conducting randomized controlled trials for these proteins remains limited. Meanwhile, observational studies can encounter several pitfalls, including uncontrolled confounding factors as well as reverse causation.

Mendelian randomization (MR) is an instrumental variable framework that can effectively mitigate biases arising from confounding and reverse causation ^17,18^. MR employs genetic variants as instruments for a risk factor (i.e. a circulating protein), and evaluates the potential causal effect of the risk factor on a disease outcome. MR relies on three core instrumental variable assumptions ^17, 18^. First, a genetic instrument should strongly predict the risk factor, known as the relevance assumption. Second, the genetic instrument should not be associated with confounders of the risk factor-disease outcome relationship, known as the independence assumption. Third, the genetic instrument should not act on the disease outcome through alternative pathways other than the instrumented risk factor, known as the no horizontal pleiotropy assumption. Recent large-scale proteo-genomic studies have identified ideal genetic instruments for circulating protein abundances, which are lead variants in cis-protein quantitative trait loci (cis-pQTLs) ^11-13^. These genetic variants are unlikely to be associated with confounders that influence the risk factor-disease outcome association, because of the randomization at conception. Moreover, their proximity to protein-coding genes suggests a direct influence on protein abundances, thereby reducing the risk of horizontal pleiotropy. Previous studies have implemented MR to investigate potential causal effects of circulating protein abundances on the risks of several complex diseases ^19-22^.

While cis-pQTL-facilitated MR can pinpoint target proteins, it is important to note that circulating proteins originate from various sources ^11-13^. These include but are not limited to endocrine cell secretion, cellular turnover and apoptosis, immune and inflammatory response, and diet and nutrition. To further understand the underpinning disease mechanisms and open new avenues for diagnostic and therapeutic advancements, it is crucial to identify candidate tissues and cell types where the target proteins are primarily produced.

In this study, we conducted integrative proteogenomic analyses to systematically identify potential biomarkers and drug targets for type 1 diabetes. We first capitalized on genetic associations from large-scale genome-wide association studies (GWASs) to conduct MR, in order to assess the associations between circulating protein abundances and type 1 diabetes risk. We then prioritized target proteins through multiple sensitivity and colocalization analyses, examinations of horizontal pleiotropy, and replications. Furthermore, we identified candidate tissues and cell types through enrichment analyses, utilizing both bulk tissue and single-cell gene expression profiles. Our findings underscore circulating proteins that exhibit a potential causal effect on the risk of type 1 diabetes.

## Results

### Target protein prioritization through Mendelian randomization

An overview of this study is presented in **Figure 1.** After identification of cis-genetic instruments and data harmonization, associations between circulating abundances of 1,565 proteins and type 1 diabetes risk were assessed using MR. MR analyses of 135 (8.6%) proteins utilized LD proxies of cis-genetic instruments **(Methods)**. Details of genetic instruments are provided in **Supplementary Table S1.**

**Figure 1.**
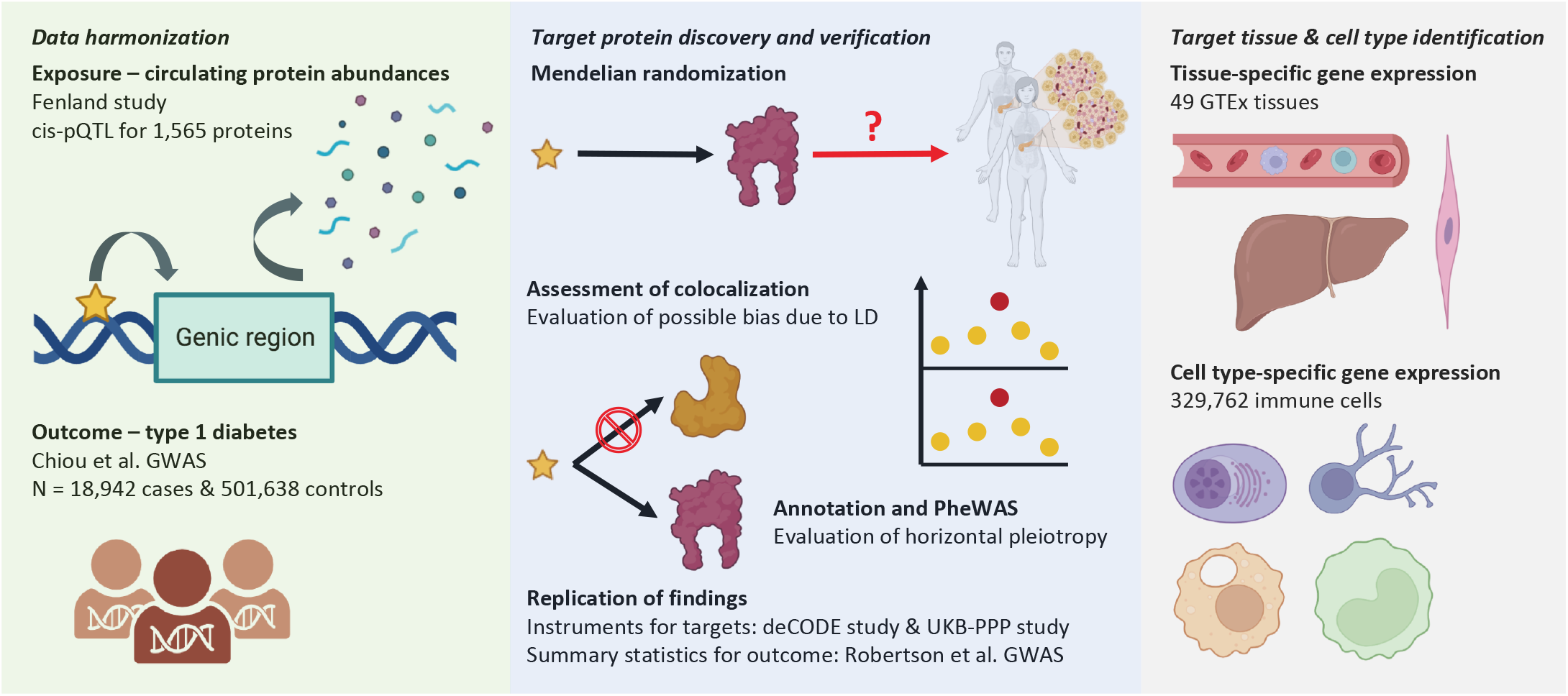
Overview of study. Mendelian randomization (MR) was conducted leveraging genome-wide association studies (GWASs) of circulating protein abundances in the Fenland study as well as a meta-analysis of type 1 diabetes GWASs. Colocalization analyses and evaluation of horizontal pleiotropy through annotation and phenome-wide association study (PheWAS) were conducted to verify MR assumptions. Significant associations were replicated using other proteomic studies and another meta-analysis of type 1 diabetes GWASs. Gene expression enrichment analyses were conducted to identify potential candidate tissues and cell types.

A total of 12 associations between circulating protein abundances and type 1 diabetes risk reached the Bonferroni-corrected significance threshold (p-value < 3.2×10^−5^; **Figure 2** and **Supplementary Figure S1**), excluding proteins whose coding genes map to the MHC region. These significant associations had a minimal F-statistic of 46.6, indicating a low risk of weak instrument bias. Full summary statistics of MR analyses are provided in **Supplementary Table S2**.

**Figure 2.**
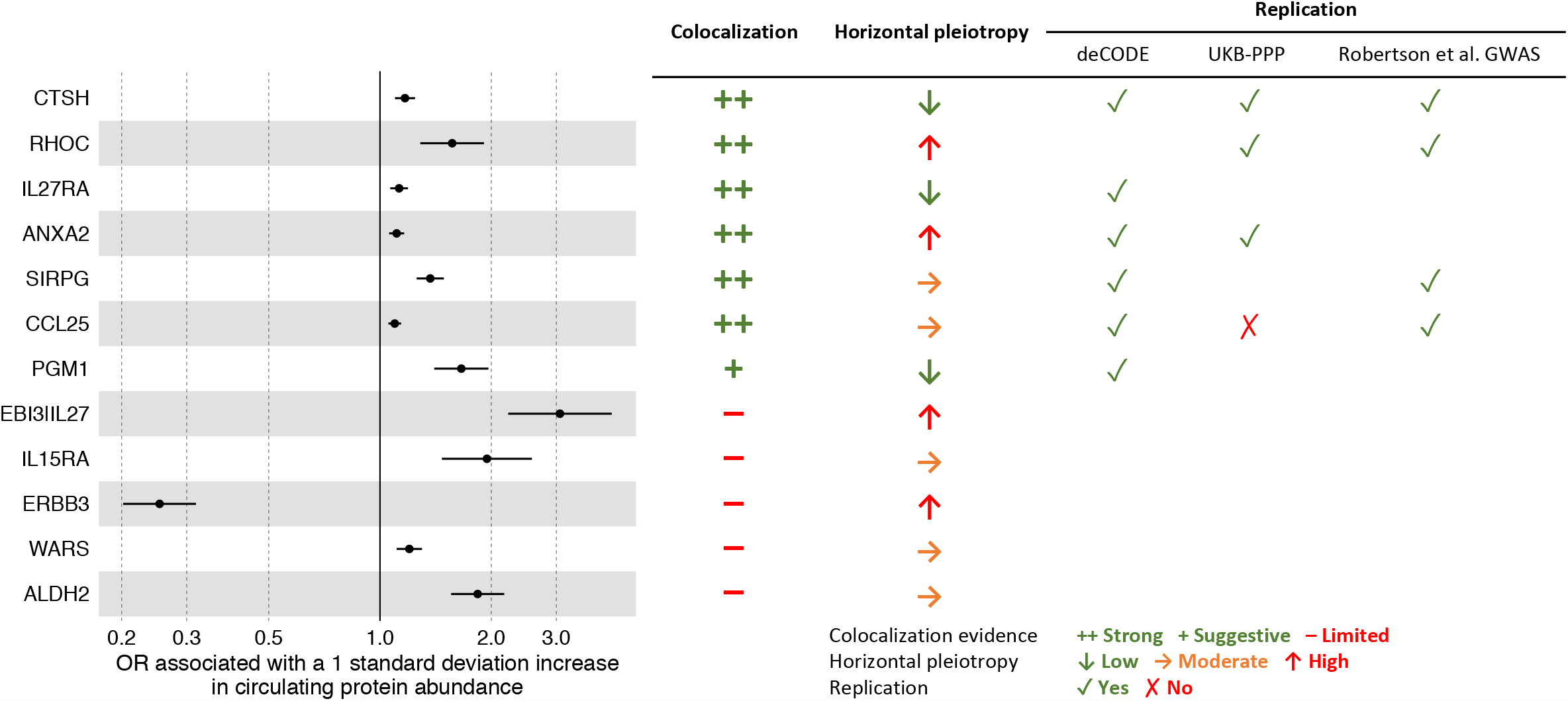
Target protein prioritization. Associations between circulating protein abundances and type 1 diabetes risk that withstood Bonferroni correction of multiple testing are illustrated. Target proteins are ordered by posterior probability of colocalization (Supplementary Table S4). A posterior probability of colocalization > 80% was considered strong evidence of colocalization, while a posterior probability of colocalization > 50% was considered suggestive evidence of colocalization. Risk of horizontal pleiotropy was assessed using V2G scores for quantifying functional connections between genetic instruments and target protein-coding genes, as well as phenome-wide associations for exploring potential pleiotropic pathways (Methods). Associations supported by strong or suggestive colocalization evidence were replicated using additional resources (Methods). Blank space indicates that no genetic instrument or proxy was identified to replicate the association. Target proteins were prioritized based on strong or suggestive colocalization evidence, the absence of a high risk of horizontal pleiotropy, and the consistent replication of associations with the risk of type 1 diabetes.

**Figure 3.**
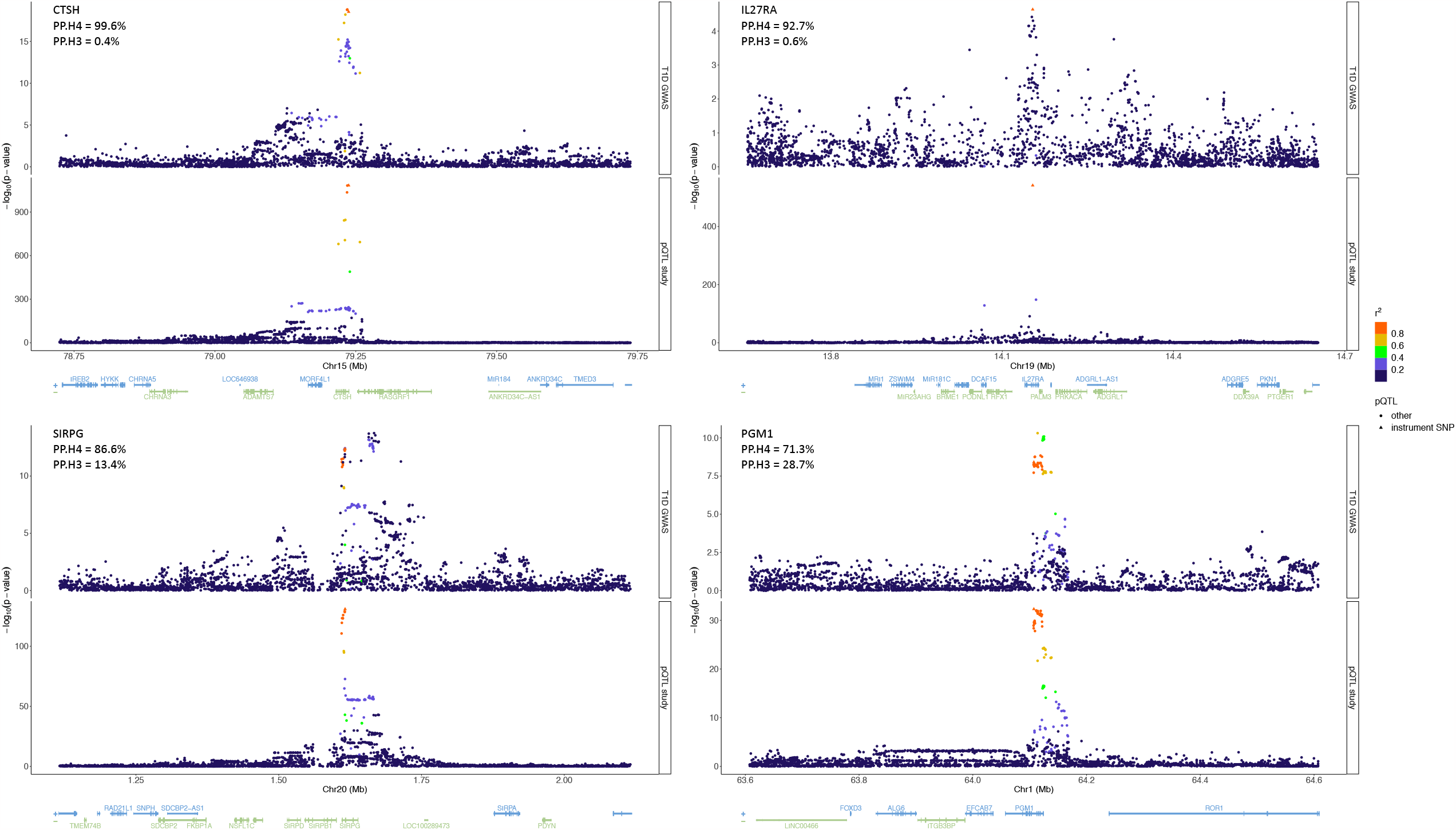
Colocalization of genetic associations with circulating abundances of prioritized target proteins and the risk of type 1 diabetes. The lead cis-genetic instruments are indicated. Genetic variants located in a ±500kb window centered around each genetic instrument are plotted with their significance in respective studies, and colored by the magnitude of correlation (linkage disequilibrium, LD r2) with the corresponding instrument. For each target protein, the posterior probability of colocalization (PP.H4) and the posterior probability of co-existence of two distinct causal variants (PP.H3) are indicated. The UCSC known gene tracks are presented, with gene models colored by their respective strands.

Of these 12 proteins, circulating abundances of CTSH, ANXA2, and CCL25 were instrumented using three cis-genetic instruments. Results of sensitivity analyses using weighted median, penalized weighted median, weighted mode, and MR-Egger methods were highly consistent with those obtained using the inverse variance weighted method **(Supplementary Table S3)**. MR-Egger intercepts largely overlapped with the null, suggesting a low risk of directional horizontal pleiotropy **(Supplementary Table S3)**.

### Colocalization evidence, horizontal pleiotropy assessment, and replication

Colocalization analyses and horizontal pleiotropy assessment were performed to verify MR assumptions for these 12 proteins **(Methods)**. Strong (PP.H4 > 80%) or suggestive (PP.H4 > 50%) evidence of colocalization between circulating protein abundance and type 1 diabetes risk was observed for CTSH, RHOC, IL27RA, ANXA2, SIRPG, CCL25, and PGM1. Conversely, colocalization evidence was limited for EBI3-IL27 complex, IL15RA, ERBB3, WARS, and ALDH2 (**Figures 2** and **3, Supplementary Figure S2**, and **Supplementary Table S4)**.

Among these target proteins supported by colocalization evidence, the genetic instruments of both RHOC and ANXA2 were predicted to have stronger functional connections to neighboring genes (i.e. *ICE2* and *ST7L*, respectively) other than their respective coding genes, as indicated by V2G scores (**Methods** and **Supplementary Table S5**). In addition, the genetic instruments of RHOC, ANXA2, SIRPG, and CCL25 have been associated with the expression or splicing of other neighboring genes, which introduces an elevated risk of horizontal pleiotropy. In contrast, the genetic instruments of CTSH, IL27RA, and PGM1 demonstrated the strongest functional connection to their respective coding genes, were not associated with the expression, splicing, or translation of other neighboring genes, and had not been associated with other known risk factors of type 1 diabetes in the Open Target database, thereby mitigating the risk of horizontal pleiotropy (**Figures 2** and **Supplementary Tables S5** and **S6**).

Seven significant associations supported by colocalization evidence were re-evaluated when cis-genetic instruments could be identified in the deCODE study or the UKB-PPP study, or when the associations could be assessed based on the type 1 diabetes GWAS meta-analysis by Robertson et al (**Methods, Figure 2,** and **Supplementary Table S7**). Six of the seven associations were replicated using these additional resources with a consistent effect direction and a similar magnitude of effect as obtained in the primary analyses **(Supplementary Table S8)**. However, based on the cis-genetic instrument identified in the UKB-PPP study, a one standard deviation increase in genetically predicted circulating abundance of CCL25 was not associated with the risk of type 1 diabetes (odds ratio, OR = 1.01; 95% CI: 0.96-1.05; p-value = 0.82; **Supplementary Table S8)**.

Following these assessments, we prioritized CTSH, IL27RA, SIRPG, and PGM1 as target proteins (**Figures 2** and **3**), while cautioning a moderate risk of horizontal pleiotropy affecting the genetic instrument of circulating SIRPG abundance. Specifically, genetically predicted circulating abundances of CTSH, IL27RA, SIRPG, and PGM1 were associated with increased odds of developing type 1 diabetes, with ORs of 1.17 (CTSH; 95%: 1.10-1.24; p-value = 9.3×10^−7^; PP.H4 = 99.6%), 1.13 (IL27RA; 95%: 1.07-1.19; p-value = 2.3×10^−5^; PP.H4 = 92.7%), 1.37 (SIRPG; 95%: 1.26-1.49; p-value = 4.3×10^−13^; PP.H4 = 86.6%), and 1.66 (PGM1; 95%: 1.40-1.96; p-value = 3.9×10^−9^; PP.H4 = 71.3%) per one standard deviation increase, respectively.

### Tissue and immune cell type enrichment of gene expression

For each of the target protein-coding genes, enrichment of gene expression in 54 tissue sites profiled by the GTEx Consortium was assessed to identify potential candidate tissues (**Methods** and **Supplementary Table S9**). As a result, the expression of *CTSH, IL27RA*, and *SIRPG* was enriched in the whole blood with a tissue-specific enrichment z-score > 10 **(Figure 4)**. Furthermore, *CTSH* expression was enriched in Epstein-Barr virus-transformed lymphocytes and SIRPG expression was enriched in the spleen, while *IL27RA* expression was enriched in both of these tissues **(Figure 4)**. In contrast, the expression of *PGM1* exhibited enrichment in skeletal muscle, heart (left ventricle), and liver **(Figure 4)**.

**Figure 4.**
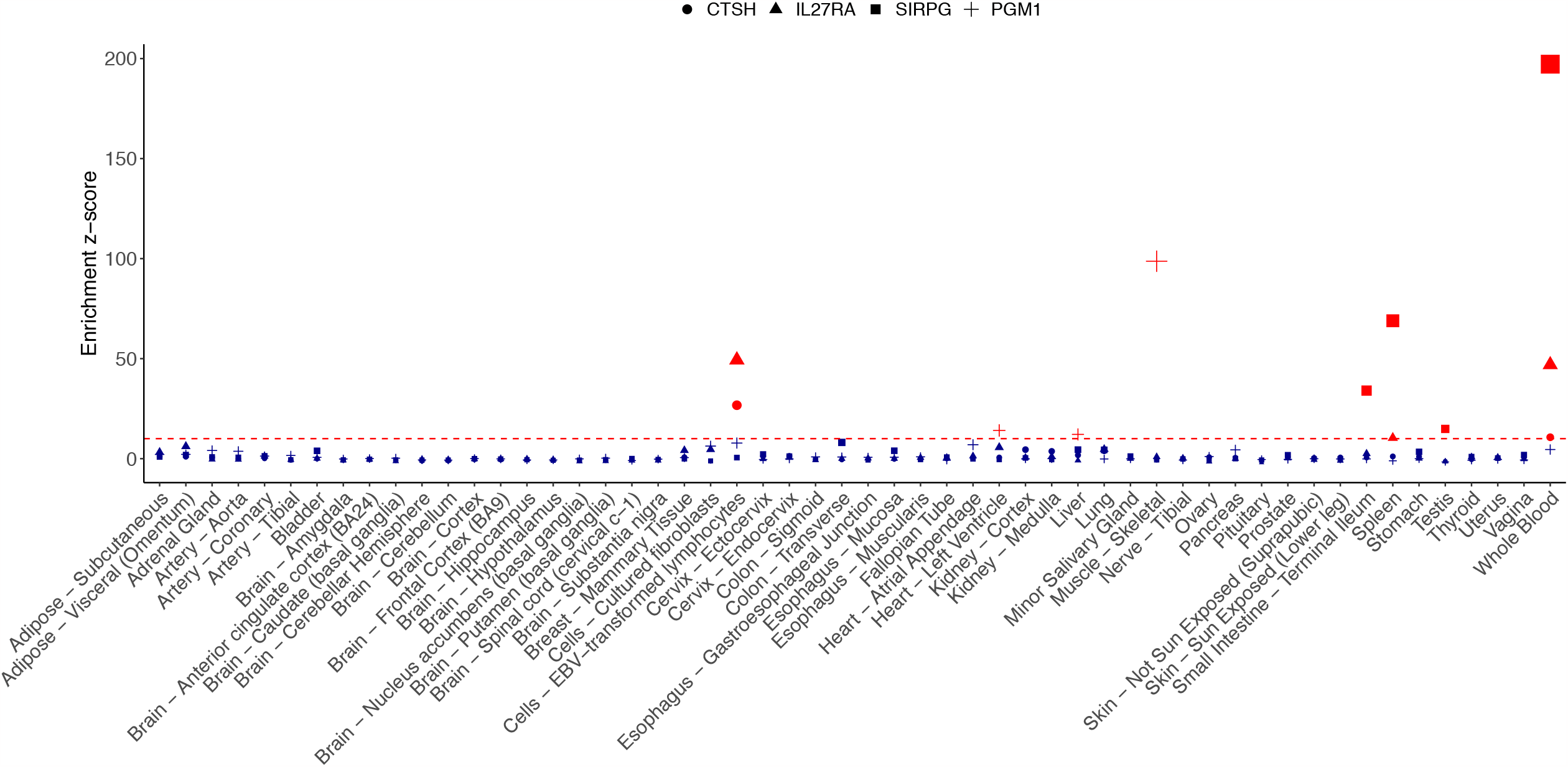
Quantification of tissue-specific gene expression. Gene expression profiles were obtained from the Genotype-Tissue Expression (GTEx) project version 8 across 54 tissue sites. Gene expression levels were normalized to account for between- and within-sample variation (Methods). For each gene, the enrichment z-scores represent standardized median gene expression levels across all tissues. Red dashed line indicates an arbitrary threshold, z-score > 10, for determining significance of enrichment.

Whole blood-specific cis-eQTL of *CTSH* and liver-specific cis-eQTL of *PGM1* demonstrated strong evidence of colocalization with the genetic associations with type 1 diabetes risk, while tissue-specific cis-eQTLs of *IL27RA* and *SIRPG* did not show evidence of colocalization (**Supplementary Figure S3** and **Supplementary Table S10**). Importantly, the cis-genetic instruments of circulating abundances of IL27RA and SIRPG were not strongly associated with their mRNA abundances in these candidate tissues **(Supplementary Figure S4)**. Meanwhile, there was strong evidence of colocalization between genetic associations with multiple isoforms of CTSH and SIRPG in whole blood and the genetic associations with the risk of type 1 diabetes **(Supplementary Table S11)**. These cis-sQTLs also overlapped with cis-pQTLs of CTSH and SIRPG **(Supplementary Figure S5)**, respectively.

Given the enrichment of *CTSH, IL27RA,* and *SIRPG* expression in immune system-related tissues, we further examined cell type-specific gene expression based on single-cell transcriptomic profiling of 329,762 immune cells, consisting of 45 curated cell types (**Methods** and **Figure 5A**). Among these immune cells, it was evident that *CTSH* expression was enriched in B cells, excluding pro-B cells and pre-B cells, as well as in myeloid cells (**Figure 5B, Supplementary Figures 6A, Supplementary Figures 7-9,** and **Supplementary Table S12**). On the other hand, *CTSH* expression was depleted in T cells, albeit with modest expression observed in effector memory CD4^+^ T cells (Teffector/EM_CD4) and tissue-resident memory T-helper 1 and T-helper 17 cells (Trm_Th1/Th17). Meanwhile, the expression level of *IL27RA* was moderate and relatively consistent across most cell types (**Figure 5C, Supplementary Figures 6B, Supplementary Figures 7-9,** and **Supplementary Table S12**). In contrast, *SIRPG* expression was enriched in T cells and natural killer cells, and depleted in B cells and myeloid cells (**Figure 5D, Supplementary Figures 6C, Supplementary Figures 7-9,** and **Supplementary Table S12**).

**Figure 5.**
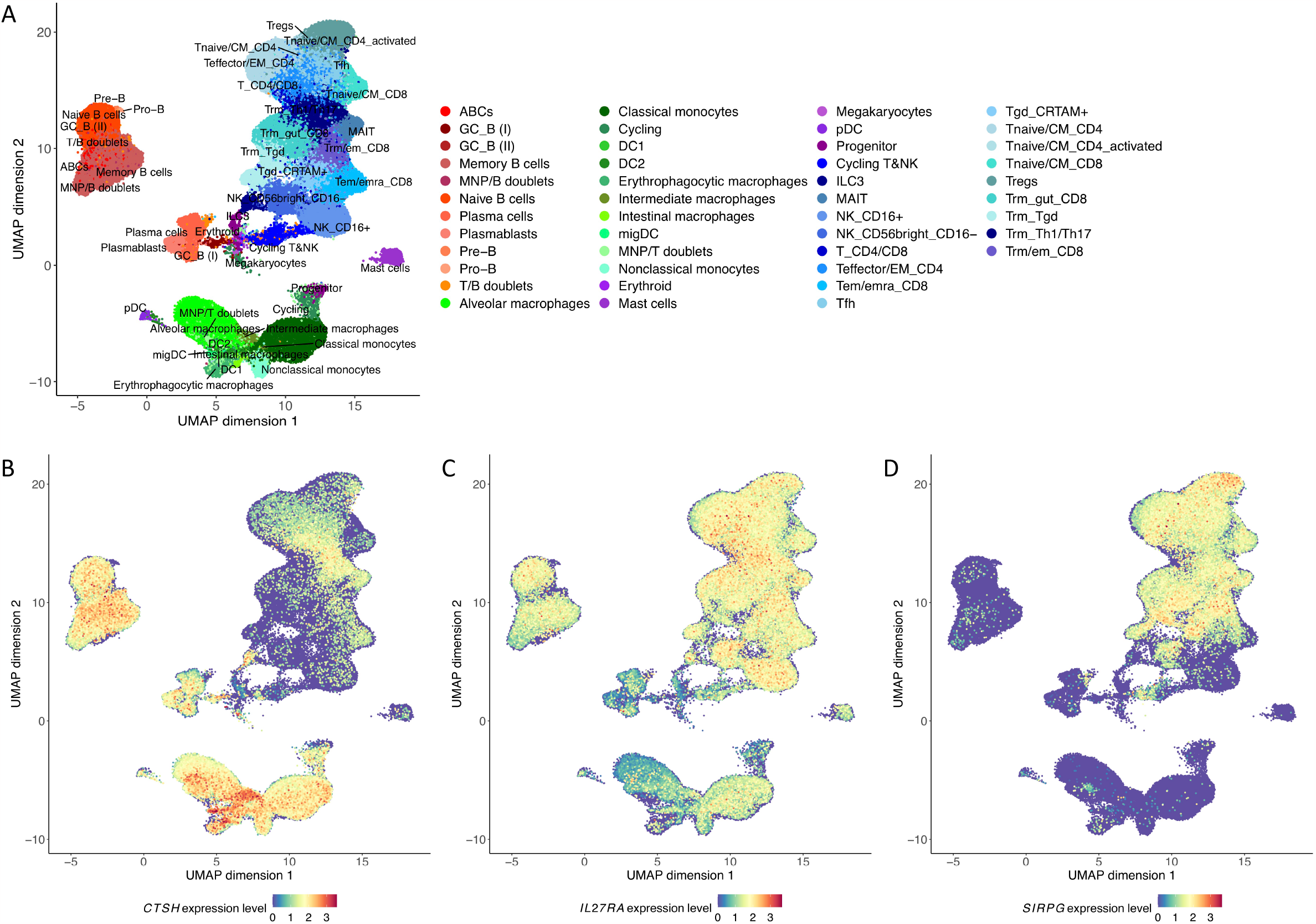
Single-cell gene expression profiles of *CTSH, IL27RA*, and *SIRPG* in immune cells. (A) Visualization of 329,762 immune cells based on Uniform Manifold Approximation and Projection (UMAP) of their transcriptomes. Cells are colored by manually curated cell types. Red colors: B cell compartment; Green colors: myeloid compartment; Purple colors: miscellaneous cell types; Blue colors: T cell compartment. Descriptions of cell types are available in Supplementary Table S11. Normalized gene expression levels of (B) *CTSH*, (C) *IL27RA*, and (D) *SIRPG* are visualized. UMAP coordinates, cell type annotations, and normalized gene expression levels were obtained from the Single Cell Portal (https://singlecell.broadinstitute.org/single_cell) under the accession ID SCP1845.

### Mendelian disorders and incident disease outcomes associated with target proteins

Among the four target proteins, PGM1 was implicated in congenital disorder of glycosylation type 1t (CDG1T), an autosomal recessive disorder caused by PGM1 deficiency due to pathogenic homozygous or compound heterozygous mutations affecting the *PGM1* gene (OMIM#614921, **Supplementary Table S13**). The other target proteins did not have known implications in Mendelian disorders.

In the UK Biobank, observational associations between circulating protein abundances and incident disease outcomes were only available for CTSH. Over 16 years of follow-up, a one standard deviation increase in circulating CTSH abundance was associated with a 1.14-fold increased hazard of mortality (95% CI: 1.10-1.17; p-value = 1.7×10^−17^), and interestingly, a 1.16-fold increased hazard of type 2 diabetes based on physician-made diagnosis (95% CI: 1.11-1.21; p-value = 4.0×10^−12^; **Supplementary Figure S10** and **Supplementary Table S14**). In addition, a one standard deviation increase in circulating CTSH abundance was associated with elevated risks of systemic lupus erythematosus (hazard ratio, HR = 1.37; 95% CI: 1.17-1.60; p-value = 1.1×10^−4^), rheumatoid arthritis (HR = 1.18; 95% CI: 1.09-1.27; p-value = 2.7×10^−5^), and chronic obstructive pulmonary disease (HR = 1.09; 95% CI: 1.05-1.14; p-value = 6.3×10^−5^; **Supplementary Figure S10** and **Supplementary Table S14**).

## Discussion

Type 1 diabetes impacts millions of individuals worldwide, causing acute and chronic complications that profoundly deteriorate the quality of life and increase mortality rates ^1-6^. Managing type 1 diabetes typically requires insulin injections for glycemic control, resulting in a significant socioeconomic burden ^23,24^. There is an urgent need for innovative strategies to prevent, intervene early, and manage the disease. In this study, we conducted MR-guided target discovery to systematically examine circulating proteins that may play a crucial role in the etiology of type 1 diabetes. We also identified candidate tissues and cell types enriched of target protein-coding gene expression. Our study presents a curated selection of candidate proteins with the potential as biomarkers or drug targets.

Our integrative proteogenomic analyses prioritized four target proteins, CTSH, IL27RA, SIRPG, and PGM1. Increased circulating abundances of these proteins were predicted to increase the risk of type 1 diabetes. Specifically, CTSH (cathepsin H) is a lysosomal cysteine protease involved in the degradation of lysosomal proteins ^25,26^. CTSH in pancreatic islets may affect *β* cell survival and insulin secretion by modulating apoptotic signaling pathways and transcription factors ^27, 28^. The genomic locus within the CTSH gene has previously been associated with the risk of type 1 diabetes ^29, 30^. In this study, we observed that *CTSH* expression was enriched in B cells and myeloid cells, implying a potential role of CTSH in antigen presentation and antibody-mediated immunity. Furthermore, although the genetic risk may be conferred by gene expression, which may be mediated by DNA methylation ^31^, colocalization between the genetic associations with multiple isoforms of *CTSH* in the whole blood and the risk of type 1 diabetes suggests that alternative splicing of *CTSH* may contribute to the disease pathogenesis. In the UK Biobank, increased circulating *CTSH* abundance was linked to higher mortality and risks of common autoimmune diseases. Importantly, while the observational association analyses did not encompass type 1 diabetes as an outcome due to the limited number of cases, increased circulating CTSH abundance was associated with an increased risk of type 2 diabetes, defined based on physician-made diagnosis. This suggests the possibility of misdiagnosing type 1 diabetes as type 2 diabetes ^3, 32, 33^ within the adult population of the UK Biobank. Increased *CTSH* expression has also been associated with early-onset type 1 diabetes and rapid decline of *β* cell function in other cohort studies ^27, 34, 35^. Taken together, our findings strongly encourage functional follow-up studies to explicate the role of CTSH in type 1 diabetes and to evaluate its potential as a biomarker or drug target.

IL27RA (alpha subunit of the interleukin 27 receptor) binds to IL27, a heterodimeric cytokine composed of IL27p28 and EBI3 subunits ^36, 37^. IL27 has both pro-inflammatory functions by mediating T-helper 1 cell differentiation and increasing interferon *γ* production ^36-38^, and anti-inflammatory functions by inhibiting pro-inflammatory cytokines in T cells and promoting the production of anti-inflammatory cytokines ^39-41^. However, due to the lack of colocalization evidence and potential horizontal pleiotropic effects, we were unable to determine the effect of IL27. On the other hand, the association between circulating abundance of IL27RA and the risk of type 1 diabetes was substantiated by multiple lines of evidence. The results of our enrichment analyses align with the involvement of IL27RA in cell-mediated and antibody-mediated immunity by mediating IL27 signaling in various immune cells ^36^. While the functions of IL27RA and IL27RA-mediated IL27 signaling in type 1 diabetes remain to be explored, we posit that IL27 and IL27RA may regulate both innate and adaptive immune responses that attack the pancreatic *β* cells.

SIRPG (signal-regulatory protein *γ*) is a receptor protein involved in the negative regulation of receptor tyrosine kinase-coupled signaling processes ^42^. It has been suggested that SIRPG signaling may play an immunoregulatory role in maintaining peripheral immune tolerance and preventing autoimmunity ^43^. In line with existing studies, our analyses demonstrated that SIRPG expression was enriched in T cells and natural killer cells, where blocking of the SIRPG-CD47 interaction has been found to inhibit superantigen-induced T cell proliferation ^42, 44, 45^. These findings imply the potential significance of investigating SIRPG as a T cell-specific target for type 1 diabetes, although it should be noted that the genetic instrument for circulating SIRPG abundance was subject to a moderate risk of horizontal pleiotropy due to its associations with the expression, splicing, or translation of neighboring genes encoding other signal-regulatory proteins, SIRPB1, SIRPB2, and SIRPD.

PGM1 (phosphoglucomutase 1) is an enzyme that catalyzes the reversible conversion between glucose 1-phosphate and glucose 6-phosphate ^46^, which are important intermediates in glucose metabolism. The Mendelian disorders of PGM1 deficiency can result in congenital disorder of glycosylation ^47^. Given the crucial functions of insulin in the uptake of glucose into cells and the regulation of glycogen synthesis and breakdown ^48^, we hypothesize that PGM1 may play a role in type 1 diabetes by affecting the balance between glycogen storage and glucose utilization, particularly in muscle and liver tissues. Notably, previous GWASs have suggested that the PGM1-increasing allele of the genetic instrument, which increases the risk of type 1 diabetes, may have a marginal protective effect against type 2 diabetes ^49, 50^, although this association was not genome-wide significant. Elucidating the precise involvement of PGM1 in diabetes mellitus necessitates further efforts.

Our study has several strengths. First, we harnessed large-scale GWASs to increase the power of MR and colocalization analyses. Specifically, we obtained genetic instruments for circulating protein abundances from a large-scale proteo-genomic study with the highest coverage of the circulating proteome to date, and conducted target discovery utilizing the largest meta-analysis of type 1 diabetes GWASs. Notably, the association between circulating SIRPG abundance and the risk of type 1 diabetes was identified in a previous MR study but was not supported by colocalization evidence ^10^, which is likely attributable to the smaller sample sizes of GWASs ^51^. This advantage is also evident when compared to GTEx cis-eQTL-based analyses, which had much smaller sample sizes and failed to demonstrate colocalization evidence in most of the candidate tissues. Second, after cis-pQTL-facilitated MR analyses, we subjected the genetic instruments to rigorous scrutiny, ensuring that the no horizontal pleiotropy assumption was not violated for prioritized proteins. Third, we bolstered the validity of our findings by replicating our results using additional resources. Importantly, analyses using the genetic instruments identified in the UKB-PPP study failed to replicate the association between circulating CCL25 abundance and type 1 diabetes risk, highlighting the potential influence of protein detection platform and study population. These analyses collectively mitigated the risk of false positive results. Although deprioritized target proteins, such as IL15RA ^52, 53^ and ERBB3 ^54, 55^, have been previously linked to the risk of type 1 diabetes in different contexts, substantiating whether these associations truly denote causal effects requires future efforts. Fourth, based upon MR-prioritized target proteins, we further identified candidate tissues and cell types where the target protein-coding gene expression was enriched. This characterization yielded valuable insights into the biological relevance, disease mechanisms, as well as the therapeutic potential of these target proteins.

Our study has important limitations. First of all, our findings have not been experimentally validated, which should be the focus of follow-up studies. Second, our analyses were restricted to populations predominantly of European ancestry. Given the substantial variability in the prevalence and strong heterogeneity of type 1 diabetes across different populations in different countries ^3-5^, it is important to exercise caution when generalizing our findings to populations of non-European ancestries. Third, it should be noted that all cis-pQTLs used in this study were identified in middle-aged and older adults, whereas the type 1 diabetes GWASs included patients who were more likely to develop the disease at a younger age. However, we posit that the cis-genetic regulation of circulating protein abundances is likely consistent across age distributions. Nevertheless, we strongly advocate for similar analyses to be conducted across populations of diverse ancestries and demographic characteristics. Fourth, although existing protein detection platforms have enabled the measurement of circulating abundances for nearly 5,000 proteins, the possibility remains that potential target proteins lack valid genetic instruments. Genetics-guided target discovery based on proteo-genomic studies featuring increased sample sizes and enhanced coverage of the circulating proteome should be pursued in the future. Last but not least, due to the strong variability and highly intricate LD structure of the MHC region, we did not prioritize MHC gene-coded proteins, despite significant associations identified through MR. Considering the well-established role of the MHC region in the pathogenesis and progression of type 1 diabetes, future efforts should be dedicated to elucidating the functional impacts of these proteins.

In conclusion, through integrative proteogenomic analyses, we identified significant associations between circulating protein abundances and the risk of type 1 diabetes, which further suggested possible causal effects of CTSH, IL27RA, SIRPG, and PGM1. The roles of CTSH, IL27RA, and SIRPG in the immune system are underscored, with enrichment of *CTSH* expression in B cells and myeloid cells, and *SIRPG* expression in T cells and natural killer cells. In contrast, PGM1 may influence the risk of type 1 diabetes through its impact on glucose metabolism, particularly in muscle and liver tissues. Exploration of these target proteins as biomarkers or viable candidates for drug targeting strategies while considering the candidate tissues and cell types should be warranted in the context of type 1 diabetes.

## Online Methods

### Genome-wide association study of circulating protein abundances

Genetic associations with circulating protein abundances were assessed in the Fenland study based on 10,708 unrelated European ancestry individuals ^12^. Details of this study have been described previously ^12, 56^. Abundances of 4,775 proteins and protein complexes from plasma samples were measured using the SomaLogic SomaScan v4 assay, which includes 4,979 distinct SOMAmer reagents. GWAS was conducted for each SOMAmer protein target, referred to as “protein” hereinafter. The circulating abundances underwent rank-based inverse normal transformation after regressing out the effects of age, sex, test site, and the first ten genetic principal components ^12^. Conditional and joint (COJO) analyses ^57^ were performed to identify conditionally independent lead variants with a p-value < 1.0×10^−11^, which represented the Bonferroni-corrected genome-wide significance threshold. Cis-pQTL variants were defined for each protein as conditionally independent lead variants located within 500 kb away from the gene body of the protein-coding gene.

### Genome-wide association study of type 1 diabetes

Genetic associations with type 1 diabetes risk were assessed in a meta-analysis of GWASs by Chiou et al. including up to 18,942 patients and 501,638 controls predominantly of European ancestry from nine cohorts ^58^. Details of the participating cohorts and the meta-analysis have been described previously ^58^. There was no known overlap between participants of the Fenland study and participants of this meta-analysis.

### Mendelian randomization and sensitivity analyses

Two-sample MR was performed based on GWAS summary statistics to test associations between the genetically predicted circulating abundance of each protein and type 1 diabetes risk. Cis-pQTL variants identified in the Fenland study were used as genetic instruments. Trans-genetic variants distal to the protein-coding genes likely act on other genes, thus to mitigate the risk of horizontal pleiotropy, they were not used. If a cis-pQTL variant was unavailable in the type 1 diabetes GWAS summary statistics, we attempted to identify a proxy as the genetic instrument using the LDlink R package ^59^. The proxy should be in high linkage disequilibrium (LD; r^2^ > 0.8) with the cis-pQTL variant based on the LD reference panel consisting of non-Finnish European ancestry populations in the 1000 Genomes Project phase 3 ^60^. GWAS summary statistics for genetic instruments were harmonized with forward strand alleles inferred using allele frequency information. Palindromic variants with high minor allele frequency (MAF > 0.42) were discarded to avoid allele mismatches.

Wald ratio estimates were derived for proteins with only one cis-genetic instrument, while inverse variance weighted estimates were derived for proteins with two or more cis-genetic instruments ^61, 62^. Associations with a p-value < 3.2×10^−5^ were considered significant, representing the Bonferroni-corrected significance threshold to account for 1,565 tests. This significance threshold may be overly conservative due to possible correlation and functional relevance between proteins, but should effectively control the false positive rate.

For significant associations where the protein abundances were instrumented using three or more cis-genetic instruments, we conducted sensitivity analyses using the weighted median, penalized weighted median, weighted mode, and MR-Egger methods ^61-64^. An association was considered robust to invalid instruments if these different methods yielded estimates with a consistent effect direction and magnitude. A significant MR-Egger intercept (p-value < 0.05) would indicate existence of directional horizontal pleiotropy ^64^. Furthermore, we calculated the F-statistic for each test, where an F-statistic < 10 would indicate a risk of weak instrument bias ^65^. MR analyses were conducted using the TwoSampleMR R package version 0.5.6 ^66^.

### Colocalization analyses

While most genetic instruments are typically not associated with confounders of the exposure-outcome relationship, MR may be confounded by LD, where two genetic variants separately influence the exposure and the outcome through different mechanisms but are correlated with each other through LD ^67^. Colocalization analyses have been widely used to assess whether the exposure and the outcome share the same causal genetic variants, in order to guard against such confounding effects ^67^.

For significant associations, we performed colocalization analyses using PWCoCo ^16^, leveraging GWAS summary statistics of all variants located within 500 kb away from the cis-genetic instruments, and an LD reference panel constructed using 5,000 randomly selected unrelated European ancestry individuals from the UK Biobank ^68^. PWCoCo builds upon the classical algorithm coloc ^69^, but allows for multiple causal variants in the same genomic region through an implementation of COJO analyses for the exposure and the outcome separately, and pairwise colocalization analyses of conditionally independent signals ^16,57^. We used default priors of PWCoCo, i.e. p_1_ (prior probability of the exposure having a causal variant) = p_2_ (prior probability of the outcome having a causal variant) = 1.0×10^−4^, and p_12_ (prior probability of the exposure and the outcome sharing the same causal variant) = 1.0×10^−5^. A colocalization probability (PP.H4) > 80% was considered strong evidence of colocalization, while a PP.H4 > 50% was considered suggestive evidence of colocalization. We excluded proteins whose coding genes map to the major histocompatibility complex (MHC) region due to the strong variability and highly intricate LD structure.

### Annotation of genetic instruments and phenome-wide association study

To further evaluate potential horizontal pleiotropic effects, we obtained variant-to-gene (V2G) annotations ^70^ and phenome-wide associations from publicly available GWASs in Open Targets ^71, 72^ (retrieved July 1, 2023) for each genetic instrument. Specifically, the V2G scores were derived from a machine learning model trained to distinguish true causal genes from neighboring genes in the same genomic region ^70^. Therefore, the V2G scores can be a quantitative measure of the functional connection between a variant and a gene.

We considered a genetic instrument to be subject to a high risk of horizontal pleiotropy if the gene with the highest V2G score paired with this variant was not the target protein-coding gene. Furthermore, a genetic variant was considered to be subject to a moderate risk of horizontal pleiotropy if it had been associated with the expression, splicing, or translation of one or more genes in proximity other than the target protein-coding gene. Additionally, variants demonstrating associations with other established risk factors of type 1 diabetes were also considered to have a moderate risk of horizontal pleiotropy.

### Replication of findings

Significant associations that were supported by strong or suggestive colocalization evidence were replicated in two ways. First, we repeated MR analyses for these proteins using cis-genetic instruments identified in the deCODE study ^13^ and the UK Biobank Pharma Proteomics Project (UKB-PPP) study ^73^. The deCODE study measured circulating plasma abundances of 4,907 SOMAmer protein targets in 35,559 individuals from Iceland, employing the same SomaLogic SomaScan v4 assay as in the Fenland study ^13^. However, the genetic architecture underlying circulating protein abundances and LD structures in the Icelandic population may differ from those in other European ancestry populations due to extensive genetic drift ^74^. On the other hand, the UKB-PPP study measured plasma circulating abundances of 2,923 protein analytes using the Olink Explore 1536 platform ^73^. The discovery of pQTLs were conducted based on 35,571 European ancestry individuals. This cohort overlapped with participants in the meta-analysis of type 1 diabetes GWASs by Chiou et al ^58^.

Second, we repeated MR analyses for these proteins leveraging a different meta-analysis of type 1 diabetes GWASs by Robertson et al. ^75^, using cis-genetic instruments identified in the Fenland study. This meta-analysis comprised up to 16,159 patients, 25,386 controls, and 6,143 trio families with an affected offspring and both parents, including 7,117 participants of non-European (African, East Asian, or admixed) ancestries ^75^. Although participants in this meta-analysis partially overlapped with those in the primary analysis by Chiou et al. ^58^, genotyping in this study by Robertson et al. was conducted using the Illumina ImmunoChip, which provided dense coverage in 188 immune-relevant genomic regions, but sparse coverage in other regions 75.

### Quantification of tissue-specific gene expression

We investigated the tissue specificity of prioritized proteins leveraging gene expression profiles from the Genotype-Tissue Expression (GTEx) project version 8 ^76^. Following previous studies ^77, 78^, we quantified the enrichment of gene expression in each of the 54 non-diseased tissue sites across approximately 1,000 individuals. Specifically, we first retained genes that were detected in at least 20% of the samples with at least 5 read counts. We performed per-tissue trimmed mean of M-values (TMM) normalization using the edgeR R package ^79^. Then, we calculated the median TMM value across individuals for each gene in each tissue. Subsequently, within each tissue, we standardized the across-individual median TMM values, using the median and the median absolute deviation across genes. Finally, for each gene, the tissue-specific enrichment z-scores were calculated by standardizing the within-tissue standardized across-individual median TMM values, using the median and the median absolute deviation across tissues. Tissues with an enrichment z-score > 10 were considered to be enriched of expression of the corresponding gene.

### Genetic effects on tissue-specific gene expression

Next, in each tissue that demonstrated enrichment of gene expression, we tested colocalization of the genetic associations with gene expression and splicing patterns and the risk of type 1 diabetes for each target protein-coding gene. Genetic associations with mRNA abundances and isoform abundances were obtained respectively from the cis-expression and cis-splicing quantitative trait loci (eQTL and sQTL) analyses conducted by the GTEx Consortium ^76^. Colocalization analyses were performed using PWCoCo with default priors as described above ^16^.

### Single-cell gene expression profiling

Since tissues that play a significant role in the immune system were implicated, we further investigated whether target protein-coding genes were enriched in specific immune cell types. We obtained single-cell gene expression profiles from a cross-tissue analysis that included high-quality 329,762 immune cells from 12 donors ^80^. Details of this study, including sample collection, single-cell RNA sequencing and paired VDJ sequencing for T cell and B cell receptors, and data processing, have been described previously ^80^. In this study, manual curation was conducted after automated annotation, using existing cell type-specific gene expression signatures to identify 45 cell types. These cell types were classified as: B cell compartment, T cell compartment (predominantly T cells and natural killer cells), and myeloid compartment (predominantly macrophages, monocytes, and dendritic cells) ^80^. We evaluated the normalized gene expression level of each target protein-coding gene in each cell and compared the distribution of gene expression levels between different cell types.

### Clinically relevant variants affecting target protein-coding genes

To assess whether the prioritized proteins may be associated with other human diseases, we queried the ClinVar database (June 9^th^, 2023) ^81^ to identify Mendelian disorders that are caused by variants affecting the target protein-coding genes. Mendelian disorder-causing variants must be pathogenic or likely pathogenic variants with at least one submitter providing assertion criteria, which should not have conflicting interpretations.

### Observational associations with incident disease outcomes in the UK Biobank

Finally, we obtained observational association test statistics from a recent study to evaluate whether measured circulating protein abundances could predict future disease outcomes in the UK Biobank over 16 years of follow-up ^82^. The associations between each protein and mortality and 23 incident morbidities were estimated using Cox proportional hazards models, based on 49,234 individuals predominantly of European ancestry, adjusted for the fixed effects of age and sex, or for age only in the case of sex-specific diseases ^82^.

## Acknowledgements

T.L. has been supported by a Schmidt AI in Science Postdoctoral Fellowship. The funder has no role in study design; collection, management, analysis and interpretation of data; or the decision to submit for publication.

## Data availability

All results generated in this study are included in Supplementary Tables.

## References

1. Gepts W. Pathologic anatomy of the pancreas in juvenile diabetes mellitus. Diabetes. Oct 1965;14(10):619–33. doi:10.2337/diab.14.10.619

2. Eisenbarth GS. Type I diabetes mellitus. A chronic autoimmune disease. N Engl J Med. May 22 22;314(21):1360–8. doi:10.1056/NEJM198605223142106

3. Atkinson MA, Eisenbarth GS, Michels AW. Type 1 diabetes. The Lancet. 2014;383(9911):69–82.

4. DiMeglio LA, Evans-Molina C, Oram RA. Type 1 diabetes. The Lancet. 2018;391(10138):2449–2462.

5. Katsarou A, Gudbjörnsdottir S, Rawshani A, et al. Type 1 diabetes mellitus. Nature reviews Disease primers. 2017;3(1):1–17.

6. Daneman D. Type 1 diabetes. The Lancet. 2006;367(9513):847–858.

7. Oram RA, Patel K, Hill A, et al. A Type 1 Diabetes Genetic Risk Score Can Aid Discrimination Between Type 1 and Type 2 Diabetes in Young Adults. Diabetes Care. Mar 2016;39(3):337–44. doi:10.2337/dc15-1111

8. Redondo MJ, Geyer S, Steck AK, et al. A Type 1 Diabetes Genetic Risk Score Predicts Progression of Islet Autoimmunity and Development of Type 1 Diabetes in Individuals at Risk. Diabetes Care. Sep 2018;41(9):1887–1894. doi:10.2337/dc18-0087

9. Sharp SA, Rich SS, Wood AR, et al. Development and standardization of an improved type 1 diabetes genetic risk score for use in newborn screening and incident diagnosis. Diabetes care. 2019;42(2):200–207.

10. Yazdanpanah N, Yazdanpanah M, Wang Y, et al. Clinically Relevant Circulating Protein Biomarkers for Type 1 Diabetes: Evidence From a Two-Sample Mendelian Randomization Study. Diabetes Care. Jan 1 1;45(1):169–177. doi:10.2337/dc21-1049

11. Sun BB, Maranville JC, Peters JE, et al. Genomic atlas of the human plasma proteome. Nature. Jun 2018;558(7708):73–79. doi:10.1038/s41586-018-0175-2

12. Pietzner M, Wheeler E, Carrasco-Zanini J, et al. Mapping the proteo-genomic convergence of human diseases. Science. Oct 14 2021:eabj1541. doi:10.1126/science.abj1541

13. Ferkingstad E, Sulem P, Atlason BA, et al. Large-scale integration of the plasma proteome with genetics and disease. Nat Genet. Dec 2021;53(12):1712–1721. doi:10.1038/s41588-021-00978-w

14. Chen Y, Lu T, Pettersson-Kymmer U, et al. Genomic atlas of the plasma metabolome prioritizes metabolites implicated in human diseases. Nat Genet. Jan 2023;55(1):44–53. doi:10.1038/s41588-022-01270-1

15. Long AE, Gillespie KM, Rokni S, Bingley PJ, Williams AJ. Rising incidence of type 1 diabetes is associated with altered immunophenotype at diagnosis. Diabetes. 2012;61(3):683–686.

16. Zheng J, Haberland V, Baird D, et al. Phenome-wide Mendelian randomization mapping the influence of the plasma proteome on complex diseases. Nature genetics. 2020;52(10):1122–1131.

17. Skrivankova VW, Richmond RC, Woolf BA, et al. Strengthening the reporting of observational studies in epidemiology using mendelian randomisation (STROBE-MR): explanation and elaboration. bmj. 2021;375

18. Skrivankova VW, Richmond RC, Woolf BA, et al. Strengthening the Reporting of Observational Studies in Epidemiology using Mendelian Randomization: the STROBE-MR Statement. JAMA. 2021;326(16):1614–1621.

19. Yao C, Chen G, Song C, et al. Genome-wide mapping of plasma protein QTLs identifies putatively causal genes and pathways for cardiovascular disease. Nat Commun. Aug 15 2018;9(1):3268. doi:10.1038/s41467-018-05512-x

20. Chong M, Sjaarda J, Pigeyre M, et al. Novel Drug Targets for Ischemic Stroke Identified Through Mendelian Randomization Analysis of the Blood Proteome. Circulation. Sep 9 2019;140(10):819–830. doi:10.1161/CIRCULATIONAHA.119.040180

21. Lu T, Forgetta V, Greenwood CMT, Zhou S, Richards JB. Circulating Proteins Influencing Psychiatric Disease: A Mendelian Randomization Study. Biol Psychiatry. Jan 1 2023;93(1):82–91. doi:10.1016/j.biopsych.2022.08.015

22. Yoshiji S, Butler-Laporte G, Lu T, et al. Proteome-wide Mendelian randomization implicates nephronectin as an actionable mediator of the effect of obesity on COVID-19 severity. Nature Metabolism. 2023;5(2):248–264.

23. Beck RW, Bergenstal RM, Laffel LM, Pickup JC. Advances in technology for management of type 1 diabetes. Lancet. Oct 5 2019;394(10205):1265–1273. doi:10.1016/S0140-6736(19)31142-0

24. Lind M, Svensson AM, Kosiborod M, et al. Glycemic control and excess mortality in type 1 diabetes. N Engl J Med. Nov 20 2014;371(21):1972–82. doi:10.1056/NEJMoa1408214

25. Conus S, Simon HU. Cathepsins: key modulators of cell death and inflammatory responses. Biochem Pharmacol. Dec 1 2008;76(11):1374–82. doi:10.1016/j.bcp.2008.07.041

26. Reiser J, Adair B, Reinheckel T. Specialized roles for cysteine cathepsins in health and disease. J Clin Invest. Oct 2010;120(10):3421–31. doi:10.1172/JCI42918

27. Floyel T, Brorsson C, Nielsen LB, et al. CTSH regulates beta-cell function and disease progression in newly diagnosed type 1 diabetes patients. Proc Natl Acad Sci U S A. Jul 15 2014;111(28):10305–10. doi:10.1073/pnas.1402571111

28. D’Angelo ME, Bird PI, Peters C, Reinheckel T, Trapani JA, Sutton VR. Cathepsin H is an additional convertase of pro-granzyme B. Journal of Biological Chemistry. 2010;285(27):20514–20519.

29. Cooper JD, Smyth DJ, Smiles AM, et al. Meta-analysis of genome-wide association study data identifies additional type 1 diabetes risk loci. Nat Genet. Dec 2008;40(12):1399–401. doi:10.1038/ng.249

30. Barrett JC, Clayton DG, Concannon P, et al. Genome-wide association study and metaanalysis find that over 40 loci affect risk of type 1 diabetes. Nature genetics. 2009;41(6):703–707.

31. Ye J, Richardson TG, McArdle WL, et al. Identification of loci where DNA methylation potentially mediates genetic risk of type 1 diabetes. J Autoimmun. Sep 2018;93:66–75. doi:10.1016/j.jaut.2018.06.005

32. Usher-Smith JA, Thompson MJ, Sharp SJ, Walter FM. Factors associated with the presence of diabetic ketoacidosis at diagnosis of diabetes in children and young adults: a systematic review. Bmj. 2011;343

33. Thomas NJ, Jones SE, Weedon MN, Shields BM, Oram RA, Hattersley AT. Frequency and phenotype of type 1 diabetes in the first six decades of life: a cross-sectional, genetically stratified survival analysis from UK Biobank. Lancet Diabetes Endocrinol. Feb 2018;6(2):122–129. doi:10.1016/S2213-8587(17)30362-5

34. Mortensen HB, Swift PG, Holl RW, et al. Multinational study in children and adolescents with newly diagnosed type 1 diabetes: association of age, ketoacidosis, HLA status, and autoantibodies on residual beta-cell function and glycemic control 12 months after diagnosis. Pediatr Diabetes. Jun 2010;11(4):218–26. doi:10.1111/j.1399-5448.2009.00566.x

35. Inshaw JR, Cutler AJ, Crouch DJ, Wicker LS, Todd JA. Genetic variants predisposing most strongly to type 1 diabetes diagnosed under age 7 years lie near candidate genes that function in the immune system and in pancreatic β-cells. Diabetes Care. 2020;43(1):169–177.

36. Yoshida H, Hunter CA. The immunobiology of interleukin-27. Annu Rev Immunol. 2015;33:417–43. doi:10.1146/annurev-immunol-032414-112134

37. Pflanz S, Timans JC, Cheung J, et al. IL-27, a heterodimeric cytokine composed of EBI3 and p28 protein, induces proliferation of naive CD4+ T cells. Immunity. Jun 2002;16(6):779–90. doi:10.1016/s1074-7613(02)00324-2

38. Owaki T, Asakawa M, Fukai F, Mizuguchi J, Yoshimoto T. IL-27 induces Th1 differentiation via p38 MAPK/T-bet- and intercellular adhesion molecule-1/LFA-1/ERK1/2-dependent pathways. J Immunol. Dec 1 2006;177(11):7579–87. doi:10.4049/jimmunol.177.11.7579

39. Artis D, Villarino A, Silverman M, et al. The IL-27 receptor (WSX-1) is an inhibitor of innate and adaptive elements of type 2 immunity. The Journal of Immunology. 2004;173(9):5626–5634.

40. Stumhofer JS, Laurence A, Wilson EH, et al. Interleukin 27 negatively regulates the development of interleukin 17-producing T helper cells during chronic inflammation of the central nervous system. Nat Immunol. Sep 2006;7(9):937–45. doi:10.1038/ni1376

41. Stumhofer JS, Tait ED, Iii WJQ, et al. A role for IL-27p28 as an antagonist of gp130-mediated signaling. Nature immunology. 2010;11(12):1119–1126.

42. Brooke G, Holbrook JD, Brown MH, Barclay AN. Human lymphocytes interact directly with CD47 through a novel member of the signal regulatory protein (SIRP) family. The Journal of Immunology. 2004;173(4):2562–2570.

43. Sharp RC, Brown ME, Shapiro MR, Posgai AL, Brusko TM. The Immunoregulatory Role of the Signal Regulatory Protein Family and CD47 Signaling Pathway in Type 1 Diabetes. Front Immunol. 2021;12:739048. doi:10.3389/fimmu.2021.739048

44. Hatherley D, Graham SC, Turner J, Harlos K, Stuart DI, Barclay AN. Paired receptor specificity explained by structures of signal regulatory proteins alone and complexed with CD47. Molecular cell. 2008;31(2):266–277.

45. Dehmani S, Nerriere-Daguin V, Neel M, et al. SIRPgamma-CD47 Interaction Positively Regulates the Activation of Human T Cells in Situation of Chronic Stimulation. Front Immunol. 2021;12:732530. doi:10.3389/fimmu.2021.732530

46. Quick CB, Fisher RA, Harris H. A kinetic study of the isozymes determined by the three human phosphoglucomutase loci PGM1, PGM2, and PGM3. Eur J Biochem. Mar 1 1;42(2):511–7. doi:10.1111/j.1432-1033.1974.tb03366.x

47. Tegtmeyer LC, Rust S, van Scherpenzeel M, et al. Multiple phenotypes in phosphoglucomutase 1 deficiency. N Engl J Med. Feb 6 2014;370(6):533–42. doi:10.1056/NEJMoa1206605

48. Wilcox G. Insulin and insulin resistance. Clinical biochemist reviews. 2005;26(2):19.

49. Inshaw JRJ, Sidore C, Cucca F, et al. Analysis of overlapping genetic association in type 1 and type 2 diabetes. Diabetologia. Jun 2021;64(6):1342–1347. doi:10.1007/s00125-021-05428-0

50. Xue A, Wu Y, Zhu Z, et al. Genome-wide association analyses identify 143 risk variants and putative regulatory mechanisms for type 2 diabetes. Nat Commun. Jul 27 2018;9(1):2941. doi:10.1038/s41467-018-04951-w

51. Forgetta V, Manousaki D, Istomine R, et al. Rare Genetic Variants of Large Effect Influence Risk of Type 1 Diabetes. Diabetes. Apr 2020;69(4):784–795. doi:10.2337/db19-0831

52. Bobbala D, Mayhue M, Menendez A, Ilangumaran S, Ramanathan S. Trans-presentation of interleukin-15 by interleukin-15 receptor alpha is dispensable for the pathogenesis of autoimmune type 1 diabetes. Cellular & molecular immunology. 2017;14(7):590–596.

53. Kuczyński S, Winiarska H, Abramczyk M, Szczawińska K, Wierusz-Wysocka B, Dworacka M. IL-15 is elevated in serum patients with type 1 diabetes mellitus. Diabetes research and clinical practice. 2005;69(3):231–236.

54. Kaur S, Mirza AH, Brorsson CA, et al. The genetic and regulatory architecture of ERBB3-type 1 diabetes susceptibility locus. Mol Cell Endocrinol. Jan 5 2005;419:83–91. doi:10.1016/j.mce.2015.10.002

55. Wang H, Jin Y, Reddy MV, et al. Genetically dependent ERBB3 expression modulates antigen presenting cell function and type 1 diabetes risk. PLoS One. Jul 26 2010;5(7):e11789. doi:10.1371/journal.pone.0011789

56. O’Connor L, Brage S, Griffin SJ, Wareham NJ, Forouhi NG. The cross-sectional association between snacking behaviour and measures of adiposity: the Fenland Study, UK. British journal of nutrition. 2015;114(8):1286–1293.

57. Yang J, Ferreira T, Morris AP, et al. Conditional and joint multiple-SNP analysis of GWAS summary statistics identifies additional variants influencing complex traits. Nat Genet. Mar 18 2012;44(4):369–75, S1-3. doi:10.1038/ng.2213

58. Chiou J, Geusz RJ, Okino M-L, et al. Interpreting type 1 diabetes risk with genetics and single-cell epigenomics. Nature. 2021;594(7863):398–402.

59. Machiela MJ, Chanock SJ. LDlink: a web-based application for exploring population-specific haplotype structure and linking correlated alleles of possible functional variants. Bioinformatics. Nov 1 2015;31(21):3555–7. doi:10.1093/bioinformatics/btv402

60. Genomes Project C, Auton A, Brooks LD, et al. A global reference for human genetic variation. Nature. Oct 1 2015;526(7571):68–74. doi:10.1038/nature15393

61. Burgess S, Small DS, Thompson SG. A review of instrumental variable estimators for Mendelian randomization. Stat Methods Med Res. Oct 2017;26(5):2333–2355. doi:10.1177/0962280215597579

62. Bowden J, Del Greco MF, Minelli C, Davey Smith G, Sheehan N, Thompson J. A framework for the investigation of pleiotropy in two-sample summary data Mendelian randomization. Stat Med. May 20 2017;36(11):1783–1802. doi:10.1002/sim.7221

63. Bowden J, Davey Smith G, Haycock PC, Burgess S. Consistent estimation in Mendelian randomization with some invalid instruments using a weighted median estimator. Genetic epidemiology. 2016;40(4):304–314.

64. Bowden J, Davey Smith G, Burgess S. Mendelian randomization with invalid instruments: effect estimation and bias detection through Egger regression. International journal of epidemiology. 2015;44(2):512–525.

65. Burgess S, Thompson SG, Collaboration CCG. Avoiding bias from weak instruments in Mendelian randomization studies. Int J Epidemiol. Jun 2011;40(3):755–64. doi:10.1093/ije/dyr036

66. Hemani G, Zheng J, Elsworth B, et al. The MR-Base platform supports systematic causal inference across the human phenome. Elife. May 30 2018;7doi:10.7554/eLife.34408

67. Zuber V, Grinberg NF, Gill D, et al. Combining evidence from Mendelian randomization and colocalization: Review and comparison of approaches. Am J Hum Genet. May 5 2022;109(5):767–782. doi:10.1016/j.ajhg.2022.04.001

68. Bycroft C, Freeman C, Petkova D, et al. The UK Biobank resource with deep phenotyping and genomic data. Nature. Oct 2018;562(7726):203–209. doi:10.1038/s41586-018-0579-z

69. Giambartolomei C, Vukcevic D, Schadt EE, et al. Bayesian test for colocalisation between pairs of genetic association studies using summary statistics. PLoS Genet. May 2014;10(5):e1004383. doi:10.1371/journal.pgen.1004383

70. Mountjoy E, Schmidt EM, Carmona M, et al. An open approach to systematically prioritize causal variants and genes at all published human GWAS trait-associated loci. Nature genetics. 2021;53(11):1527–1533.

71. Ghoussaini M, Mountjoy E, Carmona M, et al. Open Targets Genetics: systematic identification of trait-associated genes using large-scale genetics and functional genomics. Nucleic Acids Res. Jan 8 2021;49(D1):D1311–D1320. doi:10.1093/nar/gkaa840

72. Ochoa D, Hercules A, Carmona M, et al. Open Targets Platform: supporting systematic drug-target identification and prioritisation. Nucleic Acids Res. Jan 8 2021;49(D1):D1302–D1310. doi:10.1093/nar/gkaa1027

73. Sun BB, Chiou J, Traylor M, et al. Plasma proteomic associations with genetics and health in the UK Biobank. Nature. Oct 2023;622(7982):329–338. doi:10.1038/s41586-023-06592-6

74. Ebenesersdottir SS, Sandoval-Velasco M, Gunnarsdottir ED, et al. Ancient genomes from Iceland reveal the making of a human population. Science. Jun 1 2018;360(6392):1028–1032. doi:10.1126/science.aar2625

75. Robertson CC, Inshaw JRJ, Onengut-Gumuscu S, et al. Fine-mapping, trans-ancestral and genomic analyses identify causal variants, cells, genes and drug targets for type 1 diabetes. Nat Genet. Jul 2021;53(7):962–971. doi:10.1038/s41588-021-00880-5

76. Consortium GT. The GTEx Consortium atlas of genetic regulatory effects across human tissues. Science. Sep 11 2020;369(6509):1318–1330. doi:10.1126/science.aaz1776

77. Verweij N, Haas ME, Nielsen JB, et al. Germline Mutations in CIDEB and Protection against Liver Disease. N Engl J Med. Jul 28 28;387(4):332–344. doi:10.1056/NEJMoa2117872

78. Akbari P, Gilani A, Sosina O, et al. Sequencing of 640,000 exomes identifies GPR75 variants associated with protection from obesity. Science. Jul 2 2022;373(6550)doi:10.1126/science.abf8683

79. Robinson MD, McCarthy DJ, Smyth GK. edgeR: a Bioconductor package for differential expression analysis of digital gene expression data. Bioinformatics. Jan 1 1;26(1):139–40. doi:10.1093/bioinformatics/btp616

80. Dominguez Conde C, Xu C, Jarvis LB, et al. Cross-tissue immune cell analysis reveals tissue-specific features in humans. Science. May 13 2022;376(6594):eabl5197. doi:10.1126/science.abl5197

81. Landrum MJ, Lee JM, Riley GR, et al. ClinVar: public archive of relationships among sequence variation and human phenotype. Nucleic acids research. 2014;42(D1):D980–D985.

82. Gadd DA, Hillary RF, Kuncheva Z, et al. Blood protein levels predict leading incident diseases and mortality in UK Biobank. medRxiv. 2023:2023.05. 01.23288879.

