## Supplementary Figures for "Integrative proteogenomic analyses provide novel interpretations of type 1 diabetes risk loci through circulating proteins"

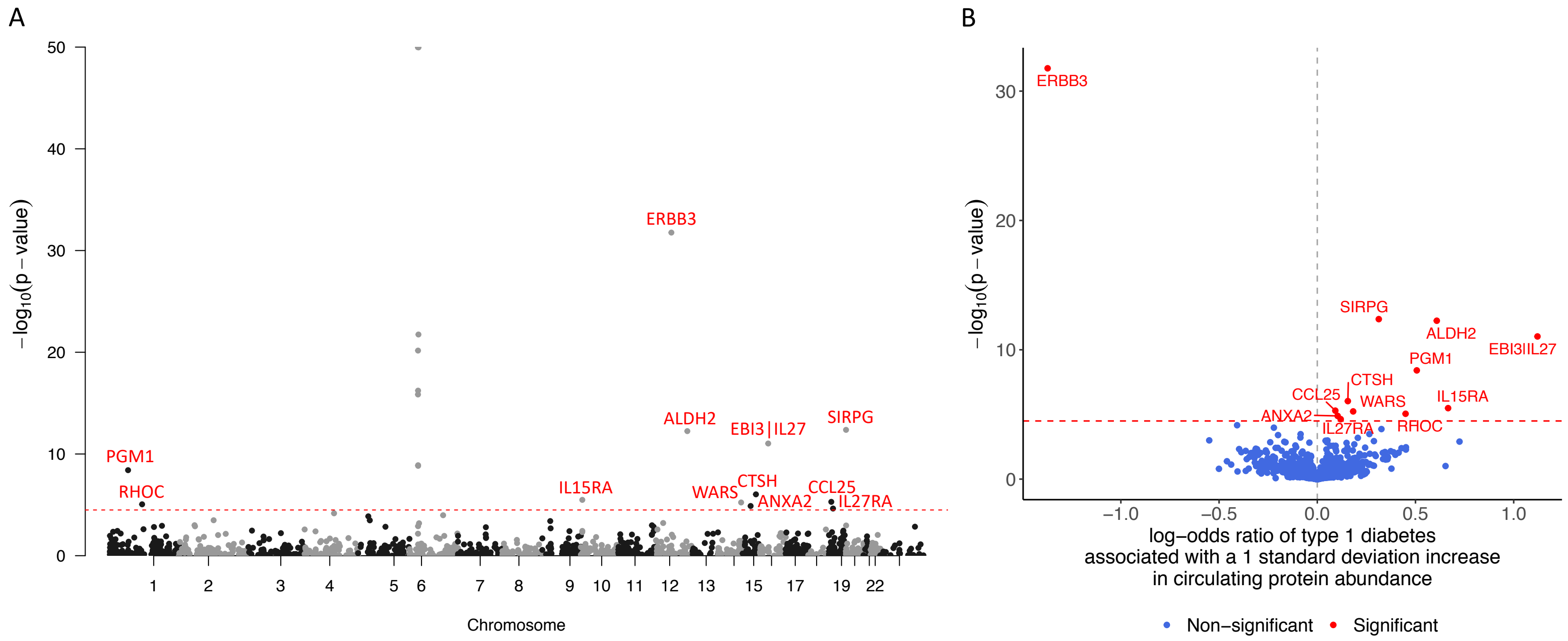


**Figure S1.** Circulating protein abundance-type 1 diabetes risk associations assessed using Mendelian randomization (MR) summarized in (A) Manhattan plot and (B) volcano plot. Each dot represents one protein. Red dashed lines denote Bonferroni-corrected significance threshold. Bonferroni-significant associations are colored red. Proteins whose coding genes map to the MHC region are not annotated in Manhattan plot and are not displayed in volcano plot. Full MR summary statistics are available in Supplementary Table S2.

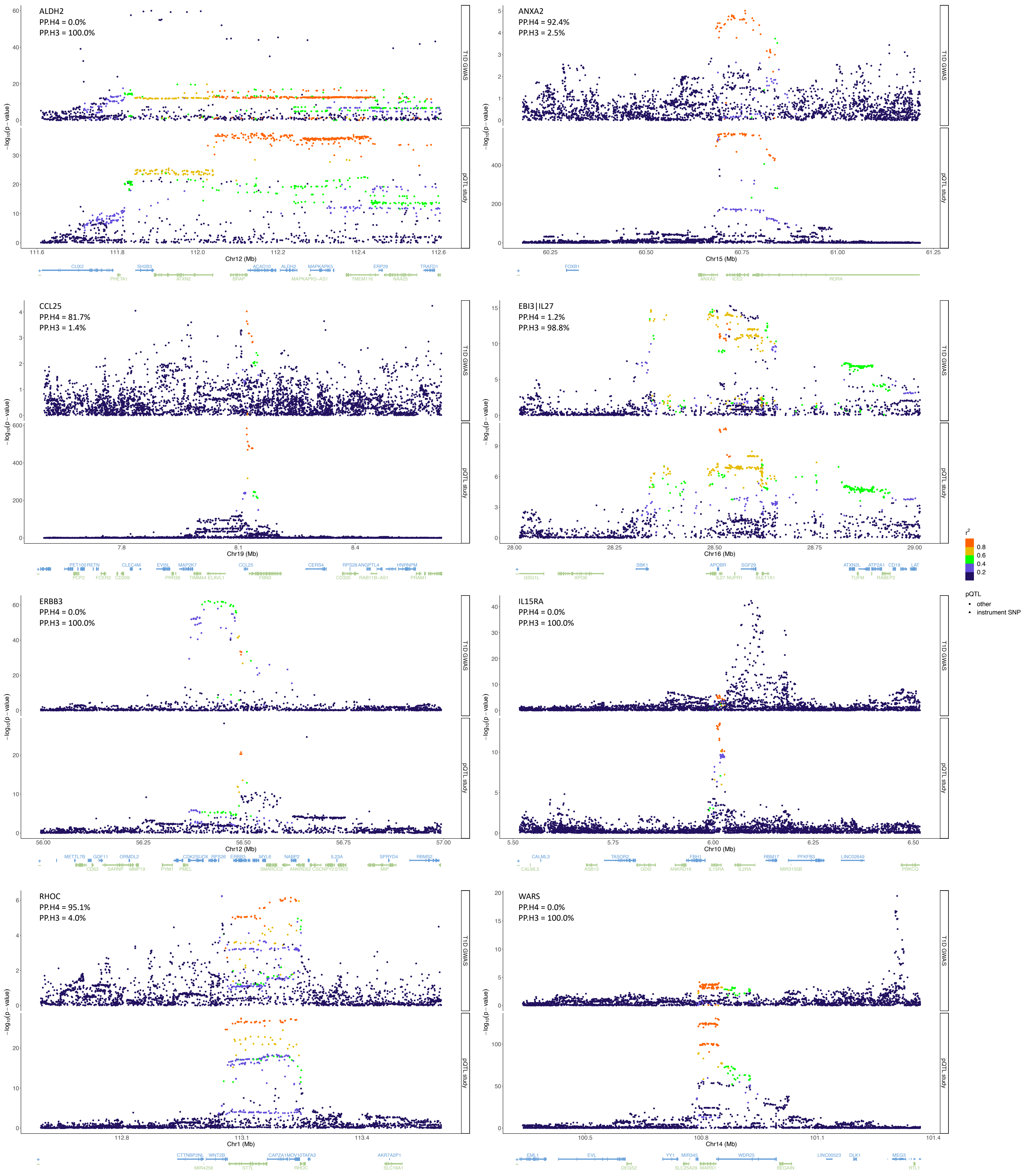


**Figure S2.** Colocalization of genetic associations with circulating abundances of deprioritized target proteins and the risk of type 1 diabetes. The lead cis-genetic instruments are indicated. Genetic variants located in a ±500kb window centered around each genetic instrument are plotted with their significance in respective studies, and colored by the magnitude of correlation (linkage disequilibrium, LD r2) with the corresponding instrument. For each target protein, the posterior probability of colocalization (PP.H4) and the posterior probability of co-existence of two distinct causal variants (PP.H3) are indicated. The UCSC known gene tracks are presented, with gene models colored by their respective strands.

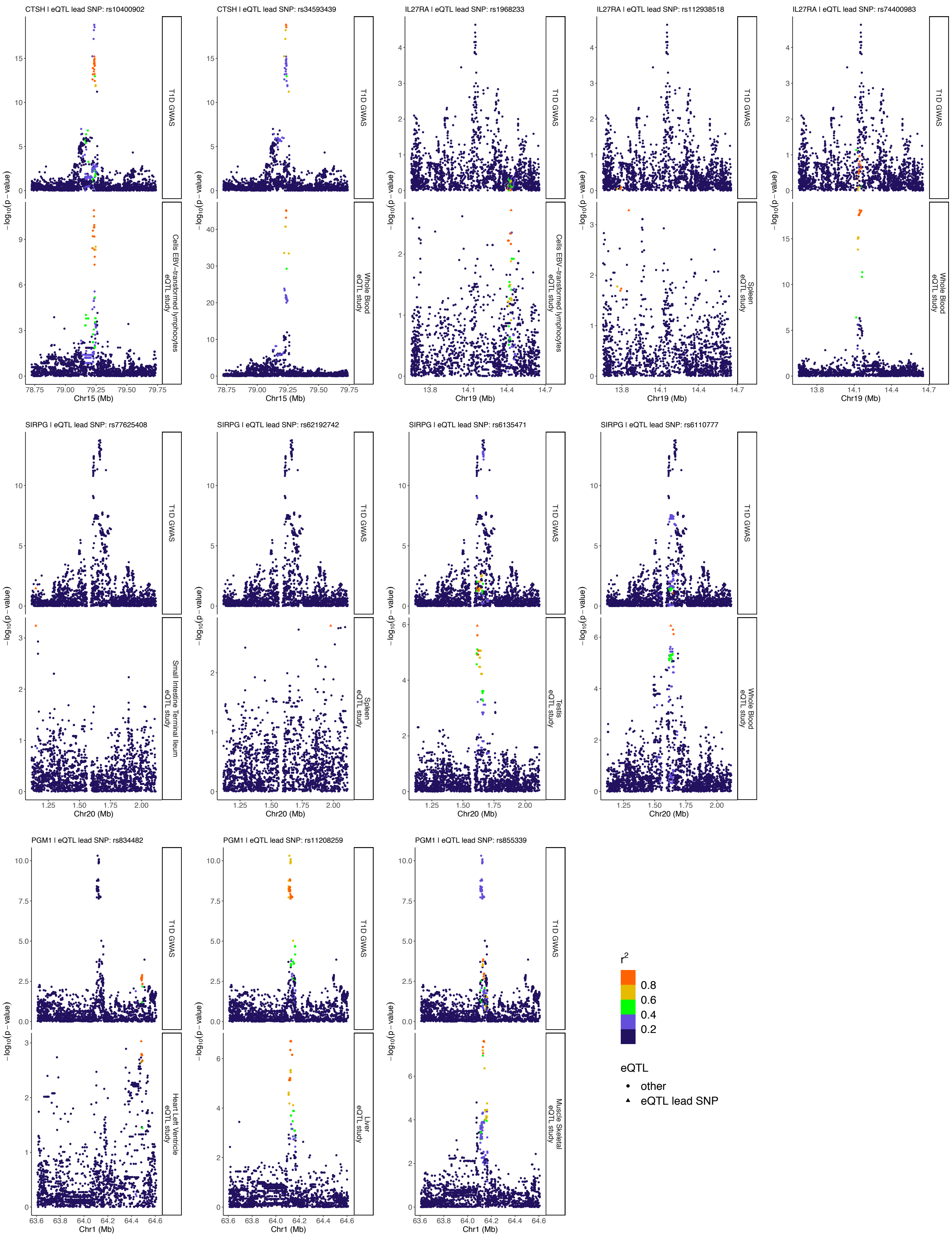


**Figure S3.** Colocalization of genetic associations with tissue-specific expression of prioritized target protein-coding genes and the risk of type 1 diabetes. The lead cis-genetic instruments of mRNA abundances are indicated. Genetic variants located in a ±500kb window centered around each genetic instrument are plotted plotted with their significance in type 1 diabetes GWAS and tissue-specific eQTL study, and colored by the magnitude of correlation (linkage disequilibrium, LD r2) with the corresponding instrument. Detailed results of colocalization analyses are provided in Supplementary Table S10.

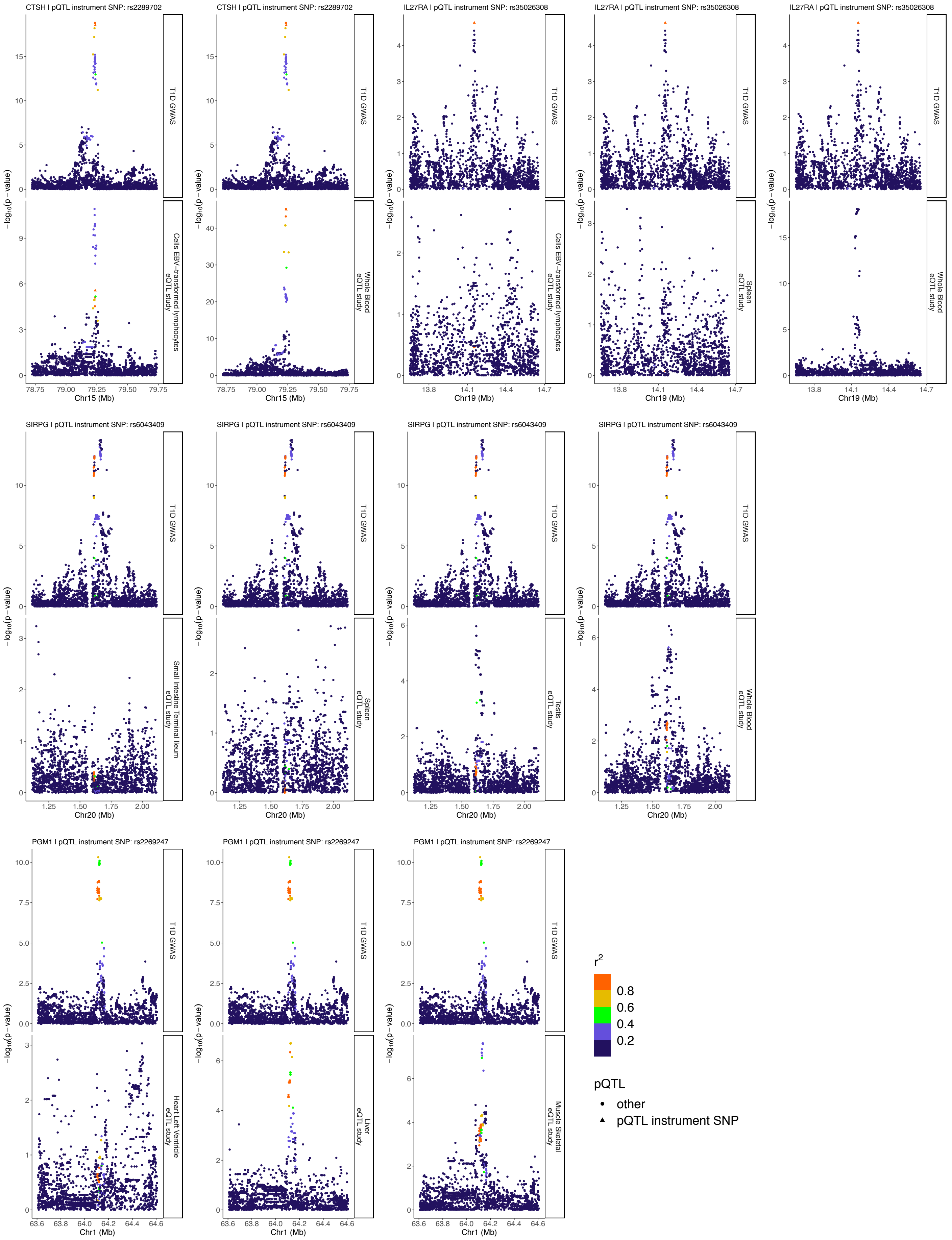


**Figure S4.** Discrepancies between cis-pQTL and cis-eQTL of prioritized target protein-coding genes. The lead cis-genetic instruments of circulating protein abundances are indicated. Genetic variants located in a ±500kb window centered around each genetic instrument are plotted with their significance in type 1 diabetes GWAS and tissue-specific eQTL study, and colored by the magnitude of correlation (linkage disequilibrium, LD r2) with the corresponding instrument.

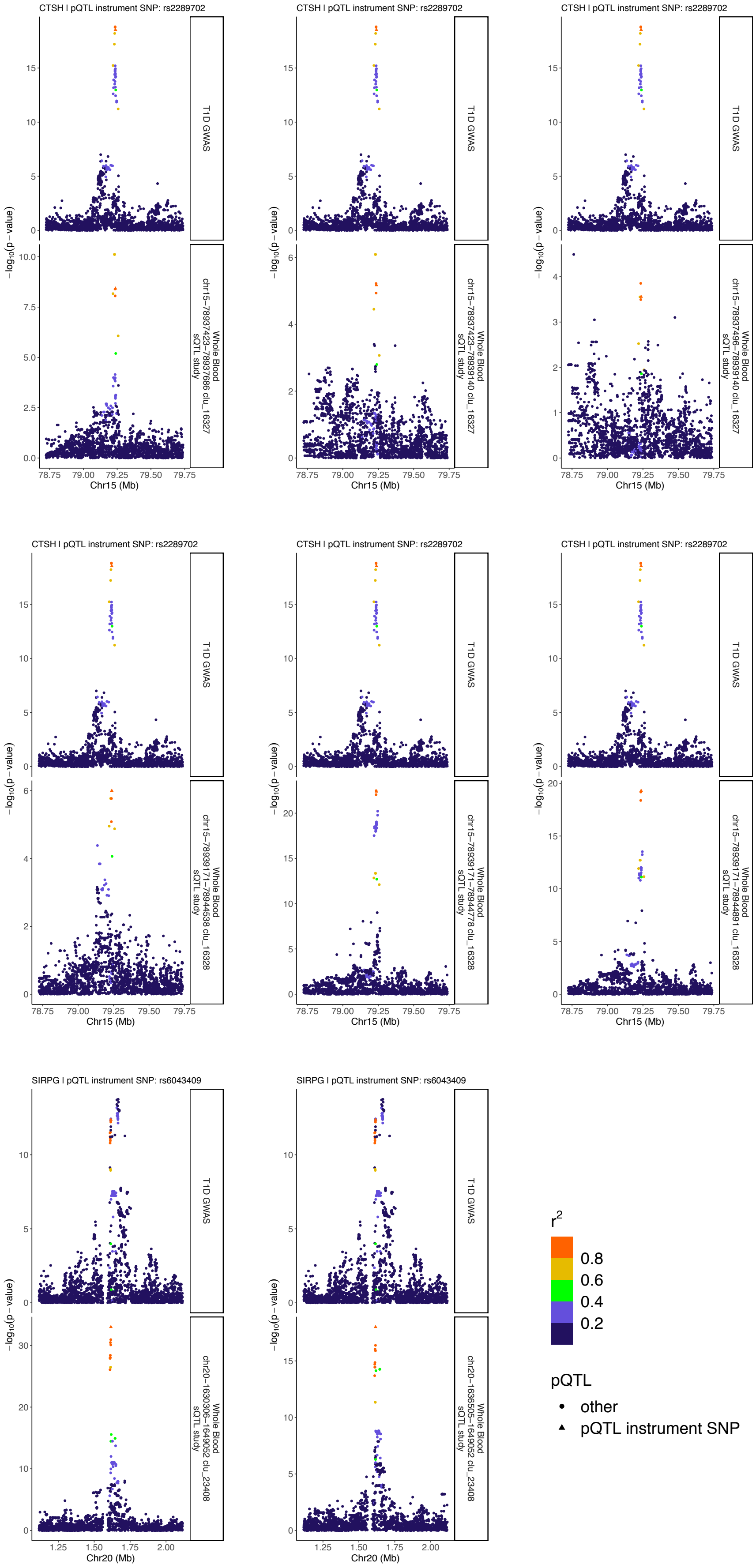


**Figure S5.** Colocalization of genetic associations with whole blood-specific alternative splicing of prioritized target protein-coding genes and the risk of type 1 diabetes. The lead cis-genetic instruments of the splicing quantitative trait loci and the corresponding intron IDs are indicated. Genetic variants located in a ±500kb window centered around each genetic instrument are plotted plotted with their significance in type 1 diabetes GWAS and whole blood-specific sQTL study, and colored by the magnitude of correlation (linkage disequilibrium, LD r2) with the corresponding instrument. Detailed results of colocalization analyses for all tissues and isoforms are provided in Supplementary Table S11. Only associations demonstrating strong evidence of colocalization are illustrated.

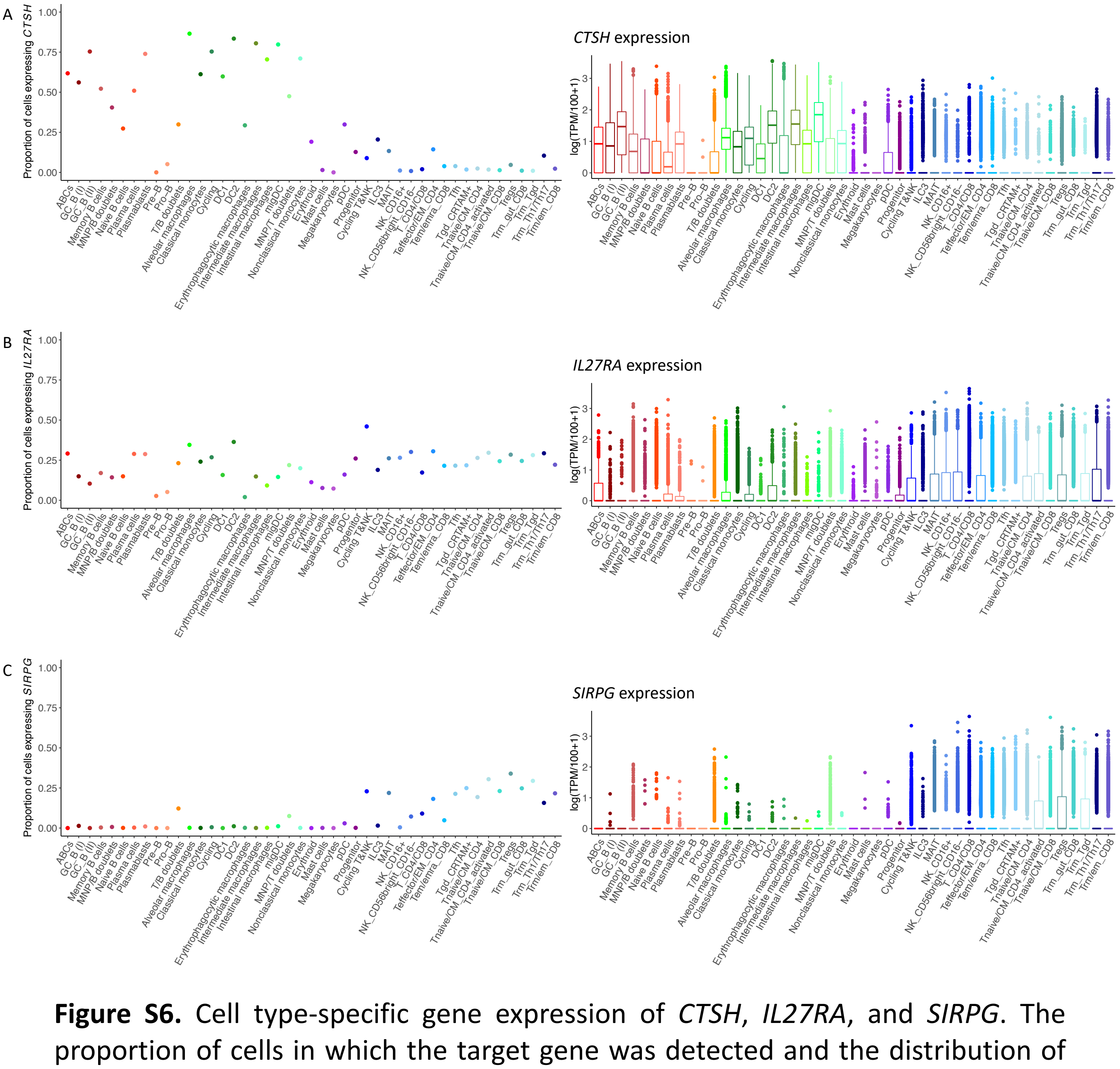


**Figure S6.** Cell type-specific gene expression of *CTSH*, *IL27RA*, and *SIRPG*. The proportion of cells in which the target gene was detected and the distribution of normalized gene expression levels in each cell type are illustrated for (A) *CTSH*, (B) *IL27RA*, and (C) *SIRPG*, colored by cell types. Descriptions of cell types are available in Supplementary Table S11. Each box represents the interquartile range (IQR) with the horizontal line inside the box denoting the median. The whiskers extend to 1.5 times the IQR from the edges of the box. Separate dots indicate outliers that fall outside the whiskers.

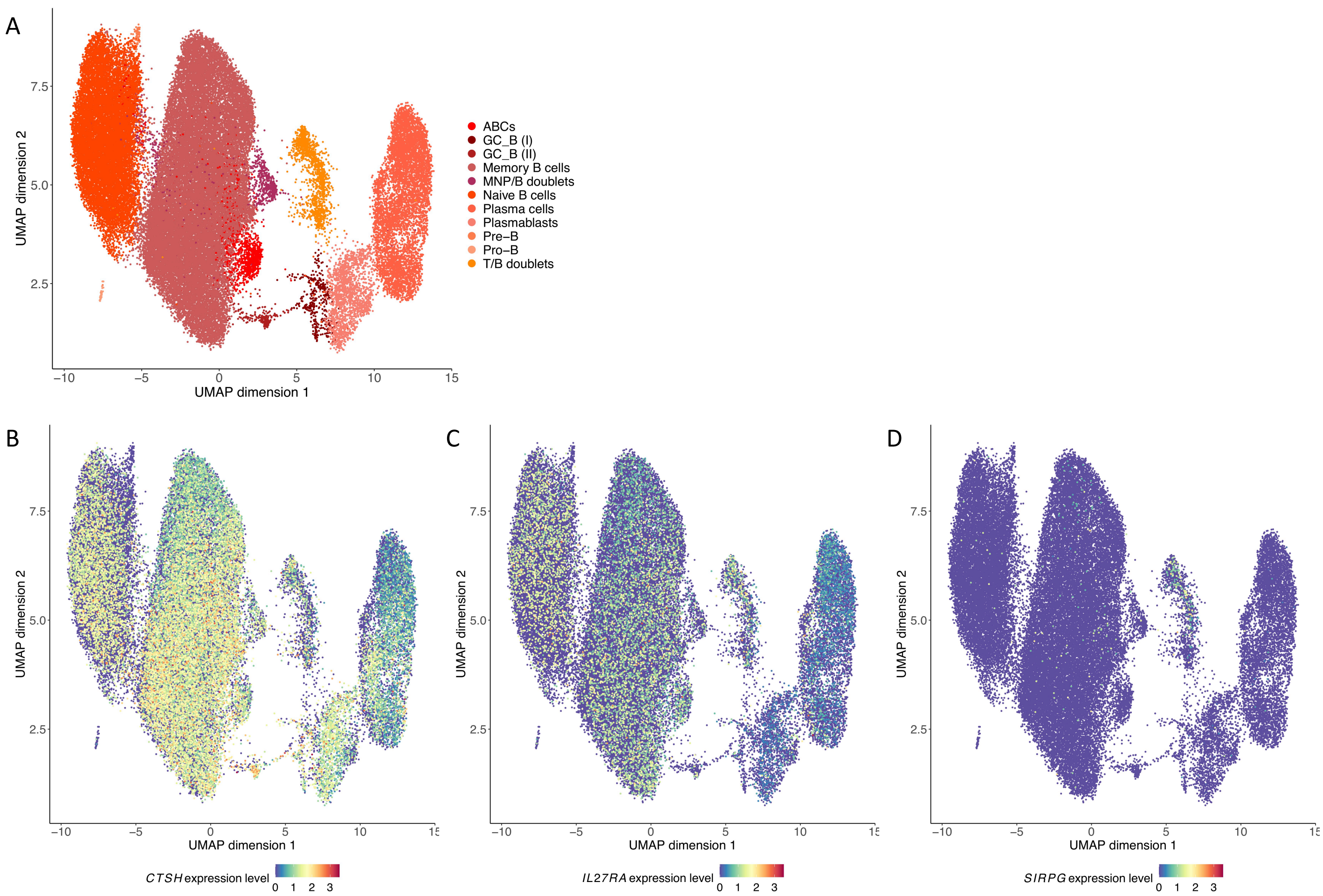


**Figure S7.** Single-cell gene expression profiles of *CTSH*, *IL27RA*, and *SIRPG* in the B cell compartment. (A) Visualization of 54,934 immune cells in the B cell compartment based on Uniform Manifold Approximation and Projection (UMAP) of their transcriptomes. Cells are colored by manually curated cell types. Descriptions of cell types are available in Supplementary Table S11. Normalized gene expression levels of (B) *CTSH*, (C) *IL27RA*, and (D) *SIRPG* are visualized. UMAP coordinates, cell type annotations, and normalized gene expression levels were obtained from the Single Cell Portal (https://singlecell.broadinstitute.org/single_cell) under the accession ID SCP1845.

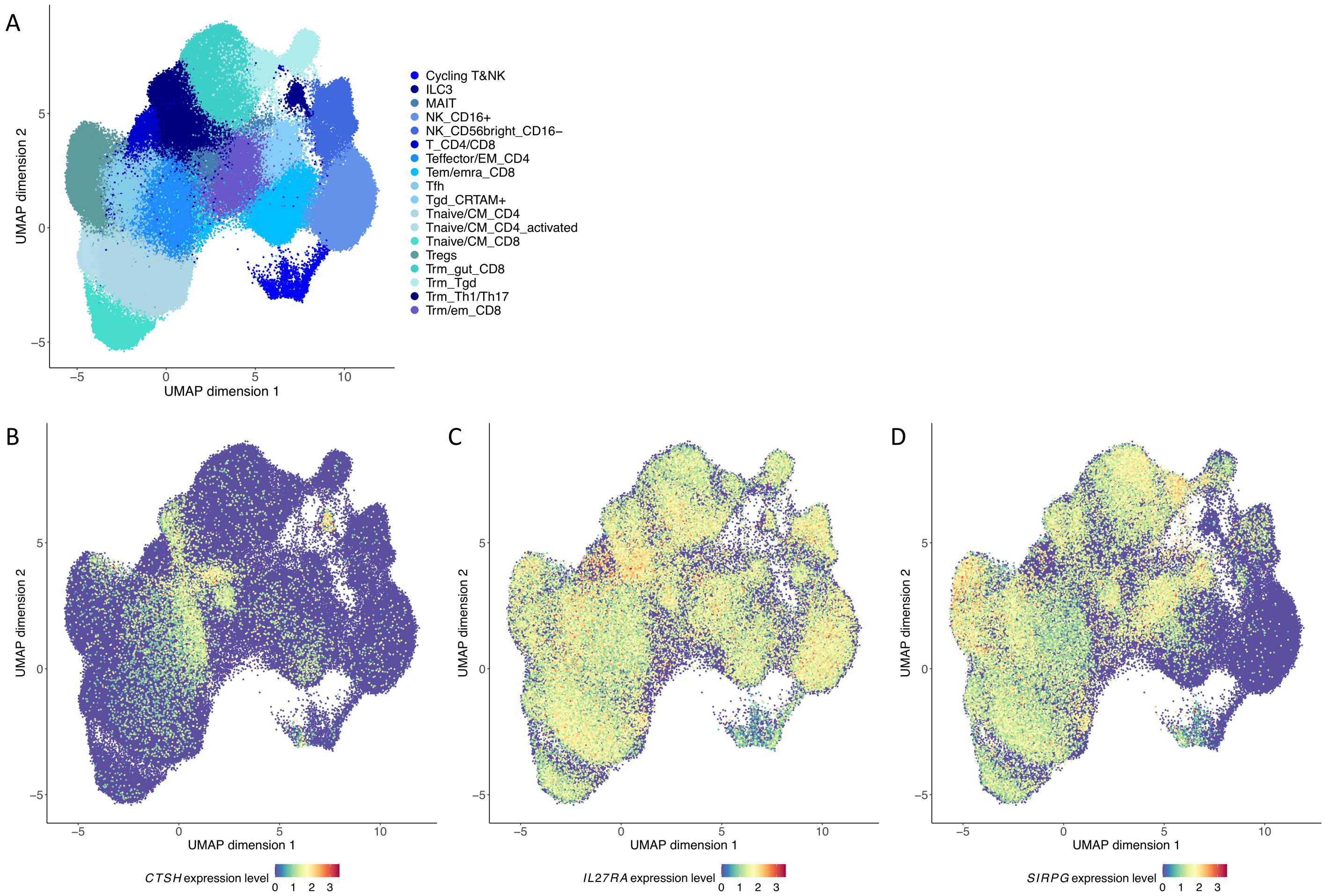


**Figure S8.** Single-cell gene expression profiles of *CTSH*, *IL27RA*, and *SIRPG* in the T cell compartment. (A) Visualization of 216,611 immune cells in the T cell compartment based on Uniform Manifold Approximation and Projection (UMAP) of their transcriptomes. Cells are colored by manually curated cell types. Descriptions of cell types are available in Supplementary Table S11. Normalized gene expression levels of (B) *CTSH*, (C) *IL27RA*, and (D) *SIRPG* are visualized. UMAP coordinates, cell type annotations, and normalized gene expression levels were obtained from the Single Cell Portal (https://singlecell.broadinstitute.org/single_cell) under the accession ID SCP1845.

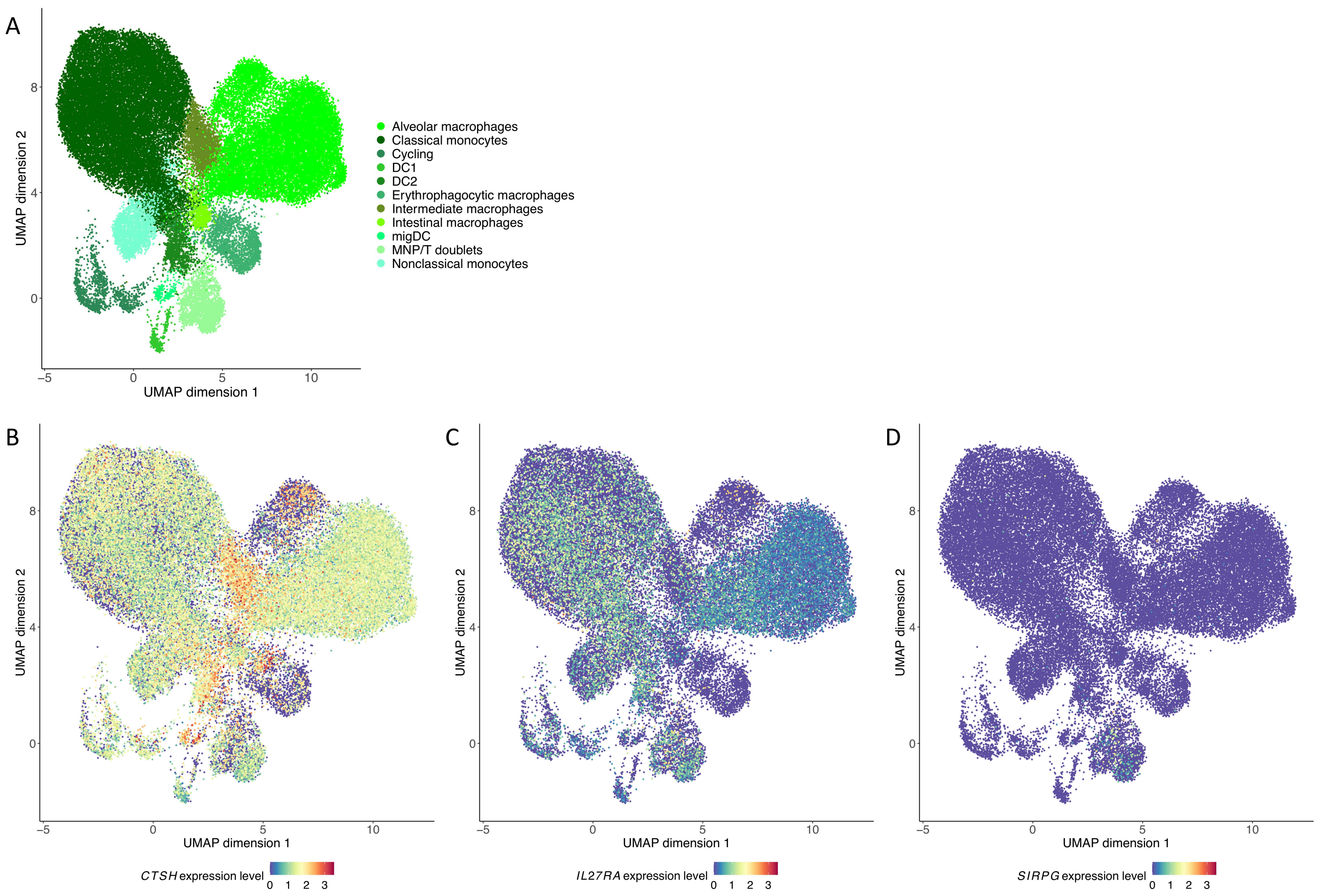


**Figure S9.** Single-cell gene expression profiles of *CTSH*, *IL27RA*, and *SIRPG* in the myeloid compartment. (A) Visualization of 51,552 immune cells in the myeloid compartment based on Uniform Manifold Approximation and Projection (UMAP) of their transcriptomes. Cells are colored by manually curated cell types. Descriptions of cell types are available in Supplementary Table S11. Normalized gene expression levels of (B) *CTSH*, (C) *IL27RA*, and (D) *SIRPG* are visualized. UMAP coordinates, cell type annotations, and normalized gene expression levels were obtained from the Single Cell Portal (https://singlecell.broadinstitute.org/single_cell) under the accession ID SCP1845.

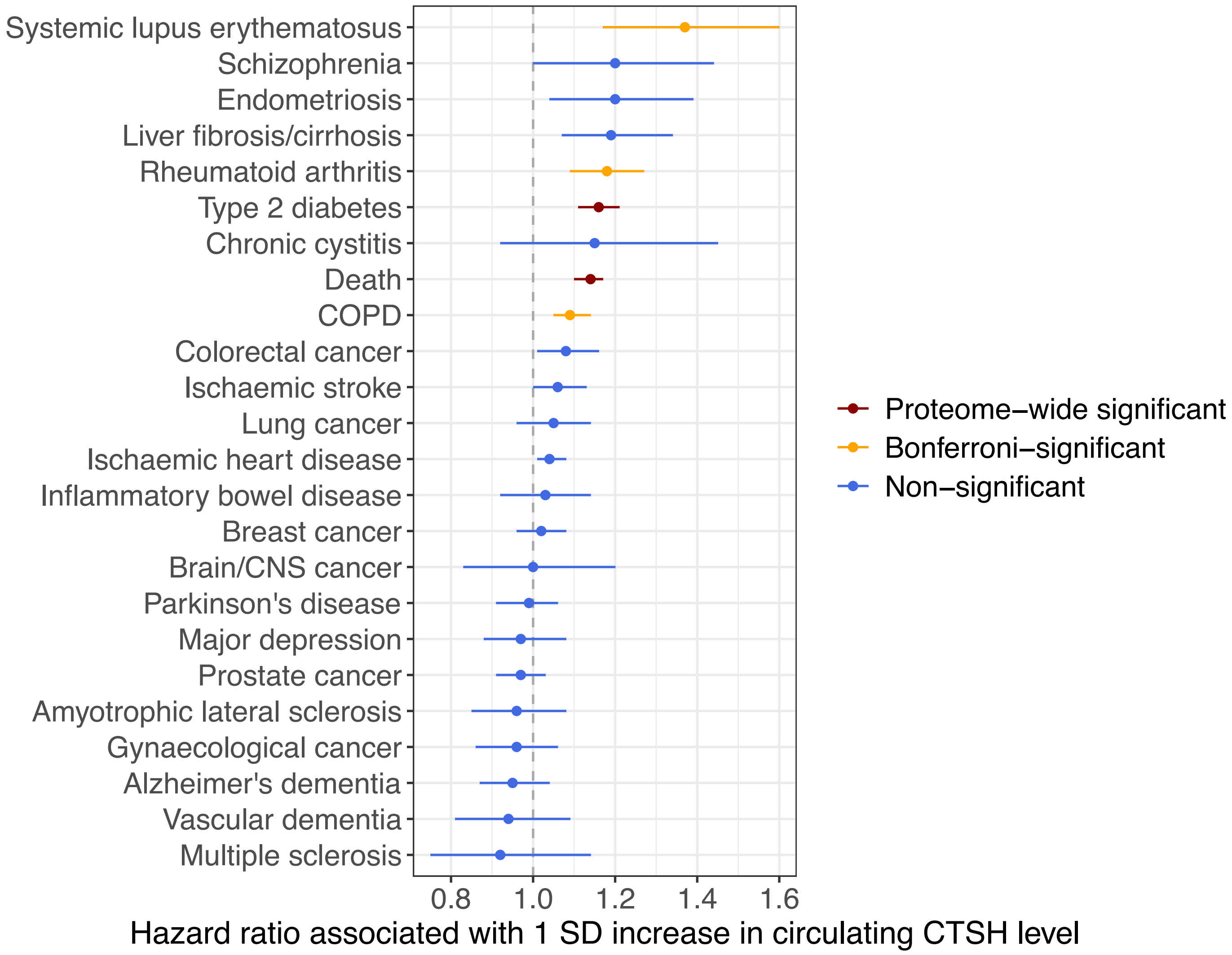


**Figure S10.** Observational associations between measured circulating CTSH abundance and incident disease outcomes in the UK Biobank. Proteome-wide significant associations accounting for the total number of tests performed across all proteins and disease outcomes (p-value < $5.4x10^{-6}$) are colored dark red. Bonferroni-significant associations accounting for 24 tests performed for CTSH (p-value < $2.1x10^{-3}$) are colored orange. Hazard ratios associated with a one standard deviation increase in circulating CTSH abundance were estimated using Cox proportional hazards models, based on 49,234 individuals predominantly of European ancestry, adjusted for the fixed effects of age and sex, or for age only in the case of sex-specific diseases.
